# Identifying Cases of Shoulder Injury Related to Vaccine Administration (SIRVA) Using Natural Language Processing

**DOI:** 10.1101/2021.05.05.21256555

**Authors:** Chengyi Zheng, Jonathan Duffy, In-Lu Amy Liu, Lina S. Sy, Ronald A. Navarro, Sunhea S. Kim, Denison Ryan, Wansu Chen, Lei Qian, Cheryl Mercado, Steven J. Jacobsen

**Author notes:** **Corresponding Author:** Chengyi Zheng, PhD, Department of Research and Evaluation, Kaiser Permanente Southern California, 100 S Los Robles Ave, 2nd Floor, Pasadena, CA 91101.

## Abstract

**Background:** Shoulder injury related to vaccine administration (SIRVA) accounts for more than half of all claims received by the National Vaccine Injury Compensation Program. However, there is a lack of population-based studies due to the challenge of identifying SIRVA cases in large health care databases.

**Objective:** To develop a natural language processing (NLP) method to identify SIRVA cases from clinical notes.

**Methods:** We conducted the study among members of a large integrated health care organization who were vaccinated between 04/1/2016 and 12/31/2017 and had subsequent diagnosis codes indicative of shoulder injury. Based on a training dataset with a chart review reference standard of 164 individuals, we developed an NLP algorithm to extract shoulder disorder information, including prior vaccination, anatomic location, temporality and causality. The algorithm identified three groups of positive SIRVA cases (definite, probable and possible) based on the strength of evidence. We compared NLP results to a chart review reference standard of 100 vaccinated individuals. We then applied the final automated NLP algorithm to a broader cohort of vaccinated individuals with a shoulder injury diagnosis code and performed manual chart confirmation on a random sample of NLP-identified definite cases and all NLP-identified probable and possible cases.

**Results:** In the validation sample, the NLP algorithm had 100% accuracy for identifying 4 SIRVA cases and 96 individuals without SIRVA. In the broader cohort of 53,585 individuals, the NLP algorithm identified 291 definite, 124 probable, and 52 possible SIRVA cases. The chart-confirmation rates for these groups were 95.3%, 67.7% and 18.9%, respectively.

**Conclusions:** The algorithm performed with high sensitivity and reasonable specificity in identifying positive SIRVA cases. The NLP algorithm can potentially be used in future population-based studies to identify this rare adverse event, avoiding labor-intensive chart review validation.

## Introduction

In 2017, shoulder injury related to vaccine administration (SIRVA) was officially added to the Vaccine Injury Table by the National Vaccine Injury Compensation Program (VICP).[1-3] The VICP defined SIRVA as shoulder pain and limited range of motion occurring after the administration of a vaccine intended for intramuscular administration in the upper arm. SIRVA is caused by an injury to the musculoskeletal structures of the shoulder (e.g., tendons, ligaments, bursae). In 2019, the number of claims related to SIRVA rose to 55% of all claims received by VICP, which resulted in an annual payout of more than $200 million.[4] Meanwhile, there has been increasing debate on whether vaccination or vaccine can cause subsequent shoulder problems.[5-7]

The debate is fueled by the lack of high-quality evidence from population-based studies. The majority of SIRVA publications are limited to case reports.[1, 8-11] Because SIRVA is a rare event but shoulder disorder is one of the most common musculoskeletal conditions, identifying SIRVA cases requires manual chart review of a large number of medical records.[11, 12]

Compared with manual chart review of medical records, natural language processing (NLP) is more efficient and produces more consistent results.[13, 14] For clinical research, NLP facilitates the identification and extraction of information that is unavailable or incomplete in structured data.[15-17] In vaccine safety studies, we have used NLP to identify two vaccine-related adverse events, anaphylaxis and local reaction.[18, 19]

### Objective

Our objective was to develop an efficient SIRVA case-finding strategy using an NLP algorithm.

## Methods

### Setting

This study was conducted at Kaiser Permanente Southern California (KPSC), an integrated health care system that provides prepaid comprehensive health care to over 4.7 million racially, ethnically, and socioeconomically diverse members.[20] KPSC’s electronic health record (EHR) system stores medical information about socio-demographics, utilization, diagnoses, laboratory tests, pharmacy utilization, membership history, and vaccination. This study was carried out using structured data and free-text clinical notes from the EHR. The Institutional Review Board at KPSC approved this study.

### Vaccinated Population With Presumptive Shoulder Injury

The study was conducted among KPSC members ≥ 3 years of age who had at least one intramuscular vaccine administered in the arm between 04/1/2016 and 12/31/2017 within a KPSC facility (Figure 1). Each vaccination was specified by the enrollee’s unique identifier, the vaccination date (index date, i.e., Day 0), and the laterality of vaccination. Membership was required for the 180 days before and after the index date.

**Figure 1.**
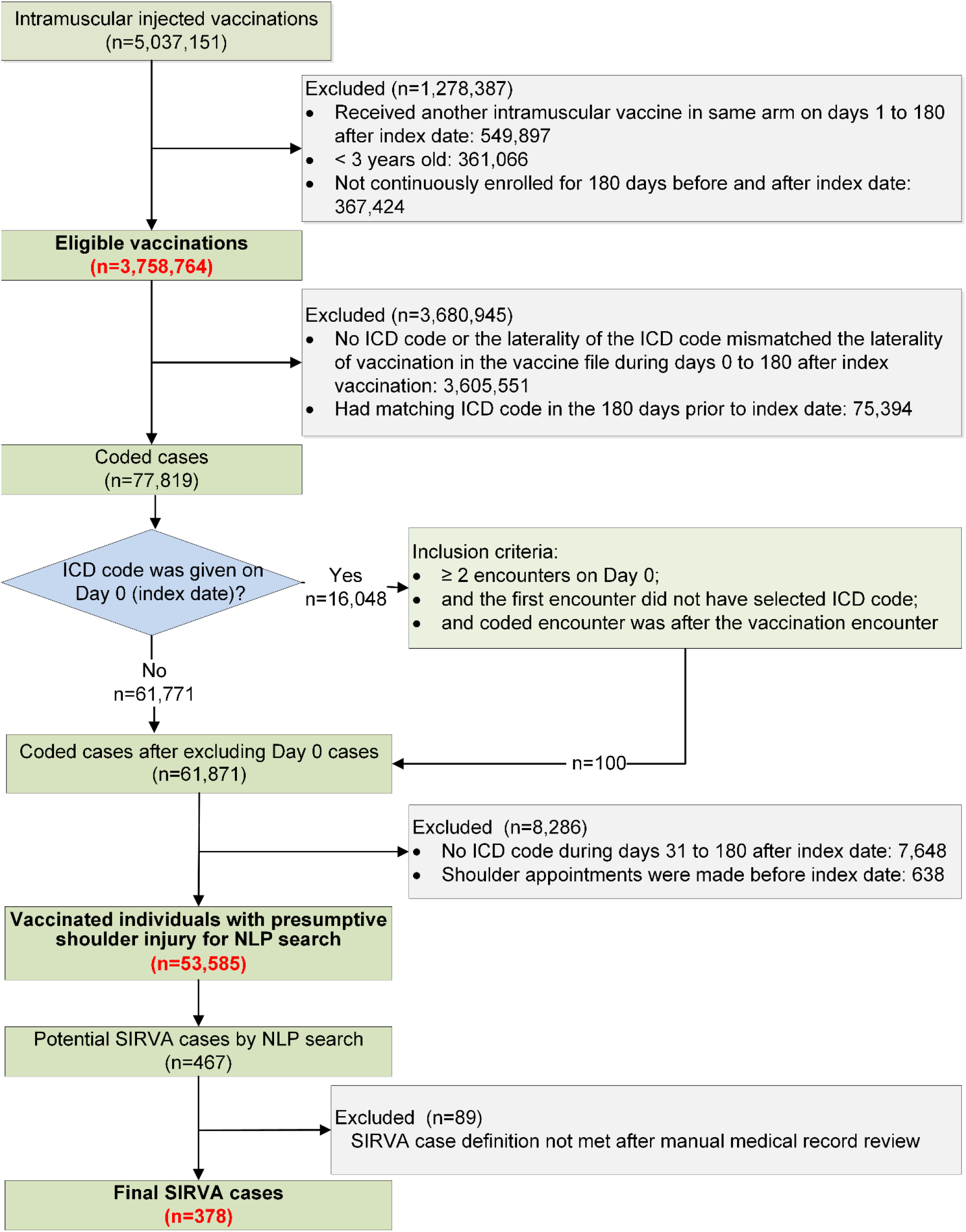
Flow Chart Showing the Selection of Eligible Vaccinated Individuals With Presumptive Shoulder Injuries, Application of The NLP Algorithm, and SIRVA Case Confirmation Results. The index date is the vaccination date. ICD (ICD-10-CM) = International Classification of Diseases, 10^th^ Revision, Clinical Modification. The complete list of ICD-10 codes is provided in Multimedia Appendix 1.

Among the vaccinated population described above, we identified individuals with a presumptive shoulder injury using International Classification of Diseases 10th Revision Clinical Modification (ICD-10-CM) codes (Multimedia Appendix 1) within 180 days after the index date; the laterality of the shoulder injury code had to match that of the vaccination. We excluded individuals who had a shoulder-related appointment or had a shoulder injury code within 180 days before the index date.

On Day 0, members could have had clinical visits with pre-existing shoulder conditions and subsequently receive vaccinations. To exclude these Day 0 pre-existing conditions, we required at least two encounters on Day 0, of which at least one of the latter encounters had to be an urgent care, emergency department, or virtual visit. Also, Day 0 encounters were excluded if the first encounter on Day 0 had a shoulder injury code or if the encounter occurred before vaccination. In order to exclude vaccine-related local reactions, one of the most common adverse events occurring shortly after vaccination, a shoulder injury code also needed to appear during Days 31-180 post-vaccination.

### SIRVA Case Definition

The VICP’s SIRVA case definition was created for medical-legal purposes. To meet this case definition, a vaccine recipient must manifest all of the following[3]:

1. Pain and reduced range of motion are limited to the shoulder in which the intramuscular vaccine was administered;
2. Pain occurs within 48 hours of vaccination;
3. No history of pain, inflammation or dysfunction of the affected shoulder prior to vaccination that would explain the alleged condition;
4. No other condition or abnormality is present that would explain the patient’s symptoms. To be eligible to file a VICP claim, symptoms must last more than 6 months after vaccination.[21]

Based on the VICP SIRVA case definition and other publications,[1, 8, 12, 22, 23] we created a SIRVA case definition suitable for a population-based study using EHR data. A valid SIRVA case needed to meet five criteria:

1. Damage to the shoulder region occurred and was confirmed by signs and symptoms (i.e., pain, limited range of motion, weakness, stiffness) and clinical diagnosis.
2. Shoulder injury occurred in the same arm in which a vaccine was injected.
3. Shoulder injury started within seven days after vaccination.
4. Vaccination was a possible cause of the shoulder injury and no other known causes were associated with the shoulder injury.
5. Shoulder injury lasted more than 30 days post-vaccination.

### Sub-population for Training and Validation of NLP Algorithm

To increase the likelihood of including true SIRVA cases in the datasets used for training and validating the NLP algorithm, we applied additional criteria to the presumptive cases to define a sub-population (n=517) (Figure 2). The criteria were as follows: 1) exclusion of individuals with an external shoulder injury (e.g., accident) code during Days -180 to 180, 2) exclusion of individuals with a shoulder injury code on Day 0, 3) requirement of a shoulder injury code during Days 1-30, and 4) requirement of a shoulder injury code on at least two different dates during Days 31-180. The criteria were based on characteristics of chart-confirmed SIRVA cases from a prior study.[12]

**Figure 2.**
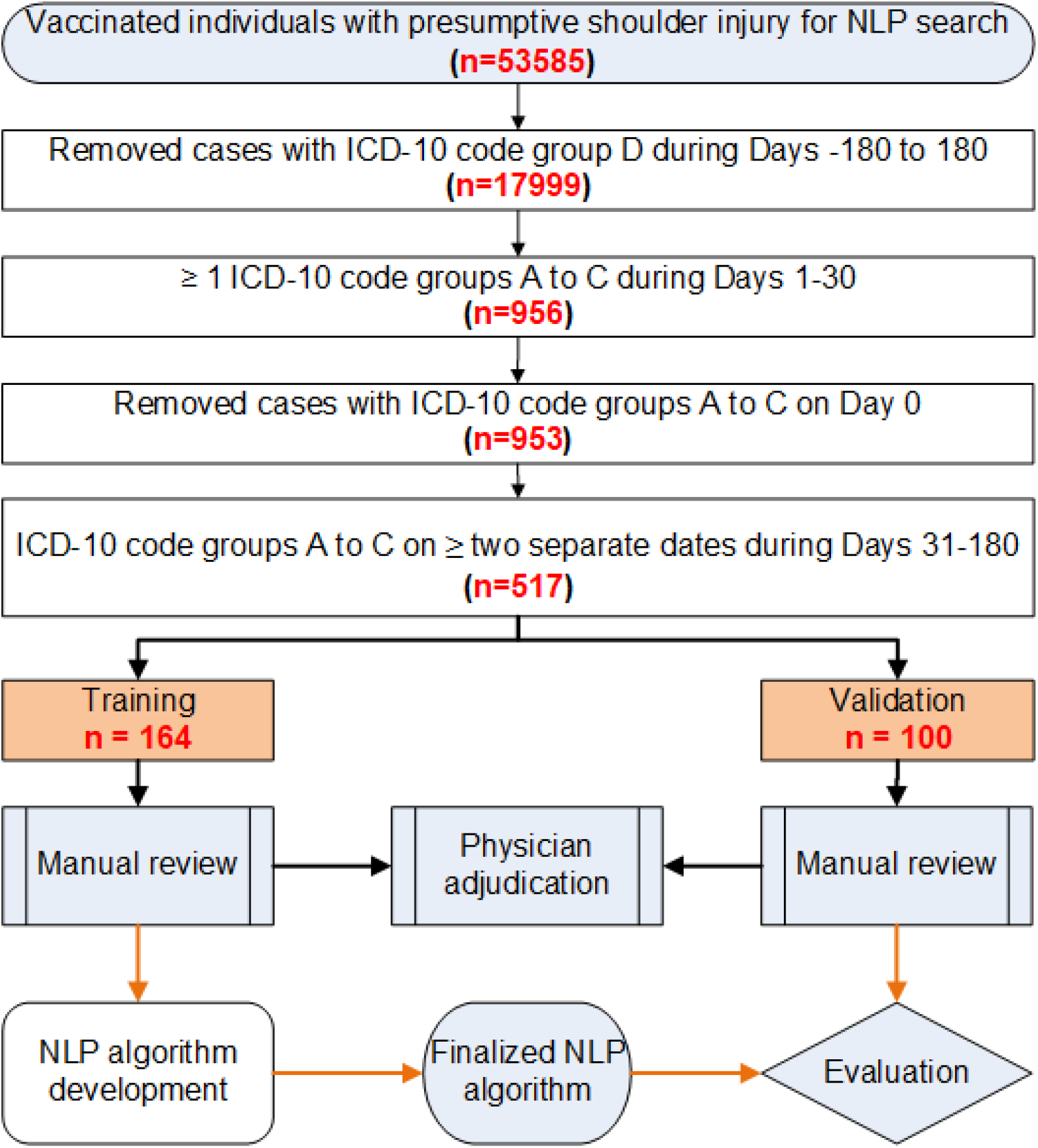
Flow Chart to Create the Dataset for Training and Validation Data Sampling ICD-10 code group A = shoulder disorder diagnoses reported in the SIRVA literature ICD-10 code group B = other shoulder disorder diagnoses not previously reported in the SIRVA literature ICD-10 code group C = shoulder symptom codes ICD-10 code group D = shoulder injury codes (ICD-10-CM chapter 19: Injury, poisoning and certain other consequences of external causes) The complete list of ICD-10 codes is provided in Multimedia Appendix 1.

#### Training Dataset (n=164)

From the sub-study population described above, we selected a random sample for chart review. The NLP algorithm was built and refined based on incremental releases of training data.[24] Once the NLP algorithm stabilized and achieved good performance, we stopped the training process. The final training dataset had 164 cases.

#### Validation Dataset (n=100)

From the remaining individuals in the sub-study population (n=353), we randomly selected another 100 cases to form the validation dataset. The chart review results were used to evaluate the performance of the final NLP algorithm.

### Manual Chart Review

We created a chart review form based on the SIRVA case definition. Chart abstractors reviewed the medical records and recorded information in the abstraction form (Multimedia Appendix 2) using the REDCap system[25]. The abstraction form was derived from a previous study of subdeltoid bursitis following vaccination, but was expanded to include other shoulder disorder diagnoses.[12] The chart abstraction and adjudication processes were similar to those used in past vaccine safety studies.[12, 19] An ascertainment period of 180 days after vaccination was used for both NLP and chart abstraction, allowing individuals sufficient time to seek medical care.[12] A second person reviewed each completed abstraction form for quality. A KPSC physician adjudicated the potential cases according to the SIRVA case definition for cases in which the chart reviewers had difficulty making a final assessment.

### NLP Terminology Development

We built customized terminologies using a set of tools, including Linguamatics I2E[26], fastText[27], Gensim[28], GloVe[29], and NLTK[30]. The terminologies were derived from various data sources, including the clinical notes of the study subjects, Vaccine Adverse Event Reporting System (VAERS) reports[31], ontologies (e.g., Unified Medical Language System (UMLS)[32]), semantic lexicons (e.g., WordNet[33]), and other online resources.

### NLP Indexing

The pre-processing steps included section detection, sentence separation, and tokenization (i.e., segmenting text into linguistic units such as words and punctuation). For each token, the indexing process added annotations for matched concepts and general linguistic entities (e.g., lexical chunks like noun or verb phrases). Additional annotations captured linguistic variations such as wildcard, substring, spelling correction, and morphological variation.

### NLP Search

We used a rule-based NLP algorithm for this study.[13, 24, 34, 35] The NLP algorithm was developed to search each indexed note at different levels: section (e.g., “Past Medical History”), intra-sentence, and cross-sentence. A distance-based relationship detection algorithm was applied to relate terms to other terms based on the number of words or sentences between them, thereby associating shoulder injury with information on vaccination site, temporality, or causality. The relationship detection algorithm also allowed for terms to be specified as ordered, or nested (e.g., an inner relation is an element of an outer relation). We used negation algorithms similar to pyConText/NegEx[36] to identify negated, uncertain and hypothetical statements. The relationship search identified the three types of information associated with shoulder injury which are described below.

#### 1. Anatomic site

The NLP algorithm extracted the body location and laterality of the shoulder injury. For example, “left” and “arm” were identified as the laterality and body location of the shoulder injury, respectively, in the sentence “Patient has persistent pain in his left arm.”

#### 2. Temporality

Temporal information was extracted by the NLP algorithm (Multimedia Appendix 3). Incomplete temporal information was inferred based on the note creation date. For example, dates with missing year information in clinical notes were assumed to occur near the note creation date. Linguistic terms, such as prepositions, were used to extract temporal relationships such as the onset date and duration linked with the vaccination event (e.g., “**for** 2 months”, “**over** the past 2 weeks”, “**since** last Thursday”).

#### 3. Causality

Within each note, the NLP algorithm searched for possible causes of shoulder injury and classified them into seven types (Table S4.1 in Multimedia Appendix 4). The determination of causal relationships between cause and shoulder injury was made by lexical-syntactic rules based on over 70 trigger terms (Table S4.2, Multimedia Appendix 4). The terminologies for causes of shoulder injury other than vaccination are listed in Table S4.3, Multimedia Appendix 4. Moreover, for each relationship search above, we also extracted the vaccine name if available because multiple vaccines could be administered concomitantly or during follow-up.

### NLP Case Classification

The final classification was based on the case definition described in the section “SIRVA case definition” by integrating vaccine, anatomic location, temporality, and causality information. Because our algorithm emphasized sensitivity, we captured additional probable and possible cases identified by NLP with weaker evidence as defined by the following three criteria:

- Cross-sentence causality: The vaccination cause was identified only by cross-sentence causal relationship search. For example, shoulder injury and vaccination were described in separate sentences:

> “Patient requesting an appointment for evaluation for left arm pain. States experiencing pain x 1 month s/p flu vaccine.”
- Vaccination was identified as a cause of shoulder injury ≤ 30 days after vaccination. Because causality was less likely to be documented when the visit date was farther away from the onset date, vaccination might only be established as the cause of the shoulder injury within ≤ 30 days of vaccination, but not > 30 days after vaccination.
- Vaccine mismatch: The vaccine associated with shoulder injury that was documented in the clinical note did not match the vaccine recorded in the vaccination file.

Positive cases which met the SIRVA case definition were further classified into three groups:

- “definite” if they met none of the three criteria;
- “probable” if they met only one of the three criteria;
- “possible” if they met ≥ 2 of the three criteria.

### NLP algorithm performance

We evaluated the NLP algorithm’s accuracy in identifying SIRVA cases compared to the chart review reference standard in the validation dataset. We calculated sensitivity, specificity, positive predictive value (PPV), and negative predictive value (NPV) and their 95% confidence intervals (CIs). Since the NLP algorithm could potentially be accurate in determining a case not to be SIRVA, but based on an incorrect assessment of an individual component of the SIRVA case definition not being met, we also conducted an error analysis of cases in which there were discrepancies between the NLP algorithm and chart review for individual components of the case definition.

### Application of NLP Algorithm to Study Population and Chart Confirmation

The final NLP algorithm was applied to the broader study population of vaccinated individuals with presumptive shoulder injury (based on codes) to identify potential SIRVA cases. We performed manual chart confirmation on a random sample of NLP-identified definite cases and all NLP-identified probable and possible cases, and calculated chart confirmation rates and their 95% CIs.

We assembled the final group of SIRVA cases based on the chart review results. We calculated the time between vaccination and the first visit for a shoulder disorder in these SIRVA cases. We also examined the vaccination-related temporal and causal statements in their clinical notes.

## Results

Out of 3,758,764 eligible vaccinations, we identified 77,819 records with a shoulder injury code (Figure 1). Among them, 16,048 had a code on Day 0. After applying the Day 0 inclusion criteria, the number of Day 0 records remaining was 100. The NLP algorithm was applied to 53,585 vaccinated individuals with presumptive shoulder injury.

### Validation Results

The NLP algorithm achieved perfect accuracy (100%) in identifying the 4 SIRVA cases from the validation dataset (n = 100). However, the small number of positive cases resulted in wide CIs for sensitivity and PPV (39.6-100.0%) (Table 1). Meanwhile, the CIs for specificity and NPV remained narrow (95.2-100.0%).

**Table 1.** Accuracy Measurements of the NLP Algorithm on Identifying SIRVA, as Compared With the Chart-confirmed Validation Data

| **Sensitivity (%)** | **Specificity (%)** | **PPV (%)** | **NPV (%)** |
| --- | --- | --- | --- |
| 100.0 | 100.0 | 100.0 | 100 |
| 39.6-100.0 | 95.2-100.0 | 39.6-100.0 | 95.2-100.0 |

Note: 95% confidence intervals are displayed beneath point estimates
PPV = positive predictive value; NPV = negative predictive value

Discrepancies between the NLP algorithm and chart review were investigated by component (Multimedia Appendix 5). For laterality, discrepancies were typically due to conflicting evidence or documentation errors in the clinical notes themselves. For temporality, the NLP algorithm incorrectly assigned symptom onset when performing cross-sentence searches, and incorrectly assigned injury duration based on incorrect laterality or capture of a resolved shoulder injury. For causality, the NLP algorithm missed causes such as daily activity and accident.

### NLP-identified Potential SIRVA Cases (n=467)

We applied the final NLP algorithm to the clinical notes of 53,585 vaccinated individuals with presumptive shoulder injury. Among them, 99.9% (n=53,530) had at least one clinical note on Days 0-180 following vaccination. The total number of clinical notes searched by NLP was 4,292,610. The average number of clinical notes per individual was 80. The NLP algorithm identified shoulder injury in 46,086 records, and 96.5% of them had matched laterality compared to the vaccination files. The NLP algorithm identified at least one cause for 55.5% of the NLP-identified shoulder injury cases. The temporal relation search identified the onset date for 99.2% of the NLP-identified shoulder injury cases. About 76% of these NLP-identified shoulder injury cases had symptom duration > 30 days post-vaccination. The number of potential SIRVA cases identified by the NLP algorithm was 467, classified into 291 definite, 124 probable, and 52 possible SIRVA cases.

### Final SIRVA Cases After Chart Review (n=378)

We performed chart review on 129 definite and all probable and possible NLP-identified SIRVA cases (Table 2). The chart confirmation rates were 95.3% [95% CI, 90.2% to 97.9%], 67.7% [95% CI, 59.1% to 75.3%], and 18.9% [95% CI, 8.7% to 30.8%] for the definite, probable, and possible groups, respectively. We did not chart review the remaining 162 NLP-identified definite cases due to a lack of resources and high confirmation rate for this group of cases. After including these 162 NLP-definite cases and 216 chart-confirmed cases, the final number of SRIVA cases was 378.

**Table 2.** Number of NLP-identified Cases and Chart-confirmed Cases

| NLP-identified group | NLP Identified | Chart Abstracted | Chart Confirmed | Confirmation Rate, % |
| --- | --- | --- | --- | --- |
| **Definite** | **291** | **129** | **123** | **95.3%** |
| **Probable** | **124** | **124** | **84** | **67.7%** |
| Cross-sentence causality | 64 | 64 | 46 | 71.9% |
| Vaccination cause identified $\leq 30$ days after vaccination | 41 | 41 | 26 | 63.4% |
| Vaccine mismatch | 19 | 19 | 12 | 63.2% |
| **Possible** | **52** | **52** | **9** | **18.9%** |
| **Total** | **467** | **305** | **216** | **70.8%** |

Among these 378 cases, the median time from vaccination to the first visit with a shoulder injury code was 43 days (interquartile range: 21–80 days, range 0-180 days). The majority of cases (361, 95.5%) had explicit temporal statements on symptoms onset in relation to vaccination. For example, “L shoulder pain that started the day she got a flu shot”, “Right shoulder pain and neck stiffness since immunizations.” The symptom onset for the remaining cases (17, 4.5%) could be derived based on the date of clinical visit, symptom duration, and causality statement. For example, “Reports having R shoulder pain for last 2 months. Thought related to vaccine she received in R arm.” There were 147 additional cases that had explicit causal statements regarding the shoulder condition and the vaccination, e.g., “status post vaccination - suspect rotator cuff irritation from vaccination itself.” Of those, 40 cases had mention of incorrect vaccine administration.

## Discussion

SIRVA is a rare outcome following vaccination without a specific diagnosis code, and it is impractical to conduct manual chart review to identify all SIRVA cases. We developed and validated an NLP algorithm to identify potential SIRVA cases with high accuracy. The only past population-based study on SIRVA[12] was limited to shoulder bursitis after influenza vaccination. In that study, a random sample of 526 out of 1098 presumptive cases was chart reviewed to identify 12 incident subdeltoid bursitis cases attributed to vaccination. In this study, we included individuals with all types of shoulder disorder diagnoses after vaccinations. Out of 53,585 presumptive cases, the NLP algorithm combined with manual chart review yielded 378 SIRVA cases. Among 3.8 million vaccinations, the rate of SIRVA in this study was around 1 per 10,000 vaccinations. It should be noted that our SIRVA case definition was different from that of the VICP and other studies in regards to symptom onset, duration, and severity.

Although the NLP algorithm’s overall accuracy was high, some challenges remained with the laterality component, despite the addition of laterality information in ICD-10-CM coding. First, descriptions of symptom location might not be precise. For example, the arm could refer to the region from the shoulder joint to the elbow joint (upper arm) or further down to the wrist. Second, the laterality recorded in the vaccine file or documented in the clinical notes could be incorrect. These issues must be considered when conducting studies using anatomic and laterality information.

There were several lessons learned from the temporality component of the NLP algorithm. First, there could be documentation of multiple onset dates during the 180 days after vaccination. Second, the disease onset information was more likely to be incomplete or inaccurate when the onset date was in the distant past. It could make it difficult to determine the onset date if the clinical visit date was further away from the vaccination date. In this study, to maximize sensitivity, any potential case with an onset falling within the predefined onset window satisfied the onset criteria.

In our study, the causality component worked reasonably well in identifying vaccination-related causality statements. While the provider or patient may have stated that the shoulder injury was vaccination-related, such statements do not provide definitive proof of causality. Because shoulder symptoms could have an insidious onset with multiple contributing factors, it was difficult to draw definitive conclusions about cause and effect. To improve specificity, we excluded cases with non-vaccination causes of shoulder injury. However, it was still challenging to identify non-vaccination causes. First, there were numerous causes of shoulder injuries. Second, some of the causes could also be the treatment for the shoulder problem. For example, exercise could be both the cause and the therapy plan for shoulder injuries. Third, the cause of shoulder injury was often not mentioned in the clinical notes. In this study, the NLP algorithm could not identify the cause in about half of the cases. Lastly, the cause of shoulder injury was often not described in the same sentence as the shoulder symptom. The cross-sentence relationship search increased the sensitivity but decreased specificity. Causal relations have been studied extensively in the NLP field [37], but only a few studies focused on health-related causal relations and were conducted using Twitter messages [38] and literature [39-41]. One study extracted causal relations from clinical text using three causal key phrases (because, due, secondary to) and discontinuation key phrases to detect adverse drug reactions in ambulatory notes and achieved high specificity (98%) but low sensitivity (31%) and PPV (45%).[42]

SIRVA-related shoulder symptoms are common for other acute or chronic medical conditions with many possible causes. Correctly integrating the NLP-identified laterality, temporality, and causality information is nontrivial. For the same patient, different clinical encounters could attribute the shoulder injury to different causes. In this study, we made patient-level classifications by utilizing the information identified from all the components from all the notes. The combination of information across multiple notes increased the sensitivity of finding SIRVA cases but reduced the specificity since the NLP algorithm could misinterpret unrelated information extracted from multiple notes.

Because we tailored the NLP algorithm to emphasize sensitivity, the confirmation rates were low in the probable (67.7%) and possible (18.9%) groups. However, since SIRVA is a rare event, manual review of all the probable and possible cases was feasible in this study. In future studies, instead of categorizing the NLP output based on the strength of evidence, one could build a machine learning model on top of the NLP outputs[13] to further improve accuracy and develop thresholds. The SIRVA cases identified in this study could also serve as training data for a machine learning algorithm.

### Limitations

This study had some potential limitations. Our study population was limited to vaccinated individuals with a diagnosis code for shoulder injury. However, loss of sensitivity is expected to be minimal since we used a comprehensive list of codes. Additionally, shoulder injuries can last a long time and are often accompanied by repeated visits. The 6-month lookback window used in this study might not have been sufficient to remove pre-existing shoulder conditions. Failure to exclude prior shoulder conditions could reduce the specificity of the NLP algorithm. In our vaccine-related local reaction study[20], most people diagnosed with a presumptive code of interest on Day 0 had symptom onset before vaccination. In this study, we excluded most cases with a shoulder injury code on Day 0. Further research is needed to study the association between SIRVA and Day 0 shoulder injury codes. Finally, because our method was tailored to this specific outcome after vaccination, its generalizability for use with other outcomes is unclear.

## Conclusion

We developed and validated an NLP algorithm to identify potential SIRVA cases among vaccinated individuals with presumptive shoulder injury. The algorithm achieved high sensitivity and reasonable specificity. The NLP algorithm can potentially be used in future population-based studies to identify this rare adverse event, avoiding labor-intensive chart review validation.

## Data Availability

The datasets generated and/or analyzed during the current study are not publicly available due to ethical standards. The authors do not have permission to share data.

## Acknowledgments

We want to thank the following persons for their contributions to data collection and medical record abstraction: Anna Lawless, Bernadine Dizon, Claire Park, Denison Ryan, Jose Pio, Joy Gelfond, Karen Schenk, Kerresa Morrissette, Melena Taylor, Nancy Canul-Jauriga, Radha Bathala, and Sunhea Kim. This study was funded through the Vaccine Safety Datalink under contract 200-2012-53580 from the Centers for Disease Control and Prevention (CDC). The findings and conclusions in this report are those of the authors and do not necessarily represent the official position of the Centers for Disease Control and Prevention.

## Abbreviations

CI: confidence interval
EHR: electronic health records
ICD-10-CM: International Classification of Diseases 10th Revision Clinical Modification
KPSC: Kaiser Permanente Southern California
NLP: natural language processing
NPV: negative predictive value
PPV: positive predictive value
SIRVA: shoulder injury related to vaccine administration
VICP: National Vaccine Injury Compensation Program

