## Supplemental material for "Identifying Cases of Shoulder Injury Related to Vaccine Administration (SIRVA) Using Natural Language Processing"

### Multimedia Appendix 1: ICD-10-CM Code Groups for Identifying Presumptive Shoulder Injury Cases

| Code group | Description |
| --- | --- |
| ICD-10 code group A | shoulder disorder diagnoses reported in the SIRVA literature |
| ICD-10 code group B | other shoulder disorder diagnoses not previously reported in the SIRVA literature |
| ICD-10 code group C | shoulder symptom codes |
| ICD-10 code group D | shoulder injury codes (ICD-10-CM chapter 19: Injury, poisoning and certain other consequences of external causes) |

ICD-10-CM = International Classification of Diseases, 10th Revision, Clinical Modification.

The complete list of ICD-10 codes for each code group is listed below.

**Table S1.1. Shoulder Disorder ICD-10-CM Codes Reported in the SIRVA Literature (ICD-10 Code Group A)**

| | | Laterality | | |
|---|---|---|---|---|
| **Code** | **Diagnosis** | **Left** | **Right** | **Unspecified** |
| **Shoulder bursitis diagnosis codes** | | | | |
| M75.5X | Bursitis of Shoulder | M75.52 | M75.51 | M75.50 |
| M71.31 | Other bursal cyst, shoulder | M71.312 | M71.311 | M71.319 |
| M71.81 | Other specified bursopathies, shoulder | M71.812 | M71.811 | M71.819 |
| **Shoulder diagnosis codes (not bursitis)** | | | | |
| M02.21X | Postimmunization arthropathy | M02.212 | M02.211 | M02.219 |
| M13.11X | Monoarthritis, not elsewhere classified, shoulder | M13.112 | M13.111 | M13.119 |
| M13.81X | Other specified arthritis, shoulder | M13.812 | M13.811 | M13.819 |
| M25.41X | Effusion of shoulder | M25.412 | M25.411 | M25.419 |
| M65.11X | Other infective tenosynovitis, shoulder | M65.112 | M65.112 | M65.119 |
| M65.81X | Other synovitis and tenosynovitis, shoulder | M65.812 | M65.811 | M65.819 |
| M67.31 | Transient synovitis, shoulder | M67.312 | M67.311 | M67.319 |
| M67.81 | Other specified disorders of synovium and tendon, shoulder | M67.812, M67.814 | M67.811, M67.813 | M67.819 |
| M67.91 | Unspecified disorder of synovium and tendon, shoulder | M67.912 | M67.911 | M67.919 |
| M67.92 | Unspecified disorder of synovium and tendon, upper arm | M67.922 | M67.921 | M67.929 |
| M75.0X | Adhesive capsulitis/frozen shoulder | M75.02 | M75.01 | M75.00 |
| M75.1XX | Tears of rotator cuff | M75.102, M75.112, M75.122 | M75.101, M75.111, M75.121 | M75.100, M75.110, M75.120 |
| M75.2X | Bicipital tendinitis | M75.22 | M75.21 | M75.20 |
| M75.3X | Calcific tendinitis of shoulder | M75.32 | M75.31 | M75.30 |
| M87.01X | Idiopathic asceptic necrosis of shoulder | M87.012 | M87.011 | M87.019 |
| M87.02X | Idiopathic asceptic necrosis of humerus | M87.022 | M87.021 | M87.029 |
| M87.11X | Osteonecrosis due to drugs, shoulder | M87.112 | M87.111 | M87.119 |
| M87.12X | Osteonecrosis due to drugs, humerus | M87.122 | M87.121 | M87.129 |
| M87.21X | Osteonecrosis due to previous trauma, shoulder | M87.212 | M87.211 | M87.219 |
| M87.22X | Osteonecrosis due to previous trauma, humerus | M87.222 | M87.221 | M87.229 |
| M87.31X | Other secondary osteonecrosis, shoulder | M87.312 | M87.311 | M87.319 |
| M87.32X | Other secondary osteonecrosis, humerus | M87.322 | M87.321 | M87.329 |
| M87.81X | Other osteonecrosis, shoulder | M87.812 | M87.811 | M87.819 |
| M87.82X | Other osteonecrosis, humerus | M87.822 | M87.821 | M87.829 |
| M89.31X | Hypertrophy of bone, shoulder | M89.312 | M89.311 | M89.319 |
| M89.51X | Osteolysis, shoulder | M89.512 | M89.511 | M89.519 |
| M89.52X | Osteolysis, upper arm | M89.522 | M89.521 | M89.529 |
| S46.00X | Unspecified injury of muscle(s) and tendon(s) of rotator cuff | S46.002* | S46.001* | S46.009* |
| S46.09X | Other injury of muscle(s) and tendon(s) of rotator cuff | S46.092* | S46.091* | S46.099* |
| S40.01X | Contusion of shoulder | S40.012* | S40.011* | S40.019* |
| S46.01X | Strain of muscle(s) and tendon(s) of the rotator cuff | S46.012* | S46.011* | S46.019* |

**Table S1.2. Shoulder Disorder ICD-10-CM Codes not Previously Reported in the SIRVA Literature (ICD-10 Code Group B)**

| | | Laterality | | |
|---|---|---|---|---|
| **Code** | **Diagnosis** | **Left** | **Right** | **Unspecified** |
| M14.61 | Charcots joint, shoulder | M14.612 | M14.611 | M14.619 |
| M21.21 | Flexion deformity, shoulder | M21.212 | M21.211 | M21.219 |
| M24.11 | Other articular cartilage disorders, shoulder | M24.112 | M24.111 | M24.119 |
| M24.21 | Disorder of ligament, shoulder | M24.212 | M24.211 | M24.219 |
| M24.41 | Recurrent dislocation, shoulder | M24.412 | M24.411 | M24.419 |
| M24.51 | Contracture, shoulder | M24.512 | M24.511 | M24.519 |
| M24.61 | Ankylosis, shoulder | M24.612 | M24.611 | M24.619 |
| M24.81 | Other specific joint derangements of shoulder, not elsewhere classified | M24.812 | M24.811 | M24.819 |
| M25.01 | Hemarthrosis, shoulder | M25.012 | M25.011 | M25.019 |
| M25.11 | Fistula, shoulder | M25.112 | M25.111 | M25.119 |
| M25.21 | Flail joint, shoulder | M25.212 | M25.211 | M25.219 |
| M25.31 | Other instability, shoulder | M25.312 | M25.311 | M25.319 |
| M25.71 | Osteophyte, shoulder | M25.712 | M25.711 | M25.719 |
| M25.81 | Other specified joint disorders, shoulder | M25.812 | M25.811 | M25.819 |
| M60.21 | Foreign body granuloma of soft tissue, not elsewhere classified, shoulder | M60.212 | M60.211 | M60.219 |
| M60.22 | Foreign body granuloma of soft tissue, not elsewhere classified, upper arm | M60.222 | M60.221 | M60.229 |
| M60.81 | Other myositis shoulder | M60.812 | M60.811 | M60.819 |
| M60.82 | Other myositis upper arm | M60.822 | M60.821 | M60.829 |
| M61.01 | Myositis ossificans traumatica, shoulder | M61.012 | M61.011 | M61.019 |
| M61.02 | Myositis ossificans traumatica, upper arm | M61.022 | M61.021 | M61.029 |
| M61.41 | Other calcification of muscle, shoulder | M61.412 | M61.411 | M61.419 |
| M61.42 | Other calcification of muscle, upper arm | M61.422 | M61.421 | M61.429 |
| M61.51 | Other ossification of muscle, shoulder | M61.512 | M61.511 | M61.519 |
| M61.52 | Other ossification of muscle, upper arm | M61.522 | M61.521 | M61.529 |
| M62.01 | Separation of muscle (nontraumatic), shoulder | M62.012 | M62.011 | M62.019 |
| M62.02 | Separation of muscle (nontraumatic), upper arm | M62.022 | M62.021 | M62.029 |
| M62.11 | Other rupture of muscle (nontraumatic), shoulder | M62.112 | M62.111 | M62.119 |
| M62.12 | Other rupture of muscle (nontraumatic), upper arm | M62.122 | M62.121 | M62.129 |
| M62.21 | Nontraumatic ischemic infarction of muscle, shoulder | M62.212 | M62.211 | M62.219 |
| M62.22 | Nontraumatic ischemic infarction of muscle, upper arm | M62.222 | M62.221 | M62.229 |
| M62.41 | Contracture of muscle, shoulder | M62.412 | M62.411 | M62.419 |
| M62.41 | Contracture of muscle, upper arm | M62.422 | M62.421 | M62.429 |
| M62.51 | Muscle wasting and atrophy, not elsewhere classified, shoulder | M62.512 | M62.511 | M62.519 |
| M62.52 | Muscle wasting and atrophy, not elsewhere classified, upper arm | M62.522 | M62.521 | M62.529 |
| M66.11 | Rupture of synovium, shoulder | M66.112 | M66.111 | M66.119 |
| M66.21 | Spontaneous rupture of extensor tendons, shoulder | M66.212 | M66.211 | M66.219 |
| M66.22 | Spontaneous rupture of extensor tendons, upper arm | M66.222 | M66.221 | M66.229 |

| | | | | |
|---|---|---|---|---|
| M66.31 | Spontaneous rupture of flexor tendons, shoulder | M66.312 | M66.311 | M66.319 |
| M66.32 | Spontaneous rupture of flexor tendons, upper arm | M66.322 | M66.321 | M66.329 |
| M66.81 | Spontaneous rupture of other tendons, shoulder | M66.812 | M66.811 | M66.819 |
| M66.82 | Spontaneous rupture of other tendons, upper arm | M66.822 | M66.821 | M66.829 |
| M67.21 | Synovial hypertrophy, not elsewhere classified, shoulder | M67.212 | M67.211 | M67.219 |
| M67.22 | Synovial hypertrophy, not elsewhere classified, upper arm | M67.222 | M67.221 | M67.229 |
| M67.41 | Ganglion, shoulder | M67.412 | M67.411 | M67.419 |
| M70.81 | Other soft tissue disorders related to use, overuse and pressure of shoulder | M70.812 | M70.811 | M70.819 |
| M70.91 | Unspecified soft tissue disorder related to use, overuse and pressure of shoulder | M70.912 | M70.911 | M70.919 |
| M75.4 | Impingement syndrome of shoulder | M75.42 | M75.41 | M75.40 |
| M75.8 | Other shoulder lesions | M75.82 | M75.81 | M75.80 |
| M75.9 | Shoulder lesion, unspecified | M75.92 | M75.91 | M75.90 |
| M79.A1 | Nontraumatic compartment syndrome of upper extremity | M79.A12 | M79.A11 | M79.A19 |
| **Other osteopathies** | | | | |
| M89.01 | Algoneurodystrophy, shoulder | M89.012 | M89.011 | M89.019 |
| M89.02 | Algoneurodystrophy, upper arm | M89.022 | M89.021 | M89.029 |
| M89.71 | Major osseous defect, shoulder region | M89.712 | M89.711 | M89.719 |
| M89.72 | Major osseous defect, humerus | M89.722 | M89.721 | M89.729 |
| M89.8X1 | Other specified disorders of bone, shoulder | | | M89.8X1 |
| M89.8X2 | Other specified disorders of bone, upper arm | | | M89.8X2 |
| **Chondropathies** | | | | |
| M94.21 | Chondromalacia, shoulder | M94.212 | M94.211 | M94.219 |
| M94.8X1 | Other specified disorders of cartilage, shoulder | | | M94.8X1 |
| M94.8X2 | Other specified disorders of cartilage, upper arm | | | M94.8X2 |
| **Other disorders of the musculoskeletal system and connective tissue** | | | | |
| M95.8 | Other specified acquired deformities of musculoskeletal system | | | M95.8 |
| M95.9 | Acquired deformity of musculoskeletal system, unspecified | | | M95.9 |

**Table S1.3. Shoulder Symptom ICD-10-CM Codes (ICD-10 Code Group C)**

| | | Laterality | | |
|---|---|---|---|---|
| **Code** | **Diagnosis** | **Left** | **Right** | **Unspecified** |
| M25.51 | Pain in shoulder | M25.512 | M25.511 | M25.519 |
| M25.61 | Stiffness of shoulder, not elsewhere classified | M25.612 | M25.611 | M25.619 |
| M79.6 | Pain in arm | M79.602 | M79.601 | M79.609 |
| M79.62 | Pain in upper arm | M79.622 | M79.621 | M79.629 |

**Table S1.4. Shoulder Injury Codes from ICD-10-CM Chapter 19: Injury, Poisoning and Certain Other Consequences of External Causes (ICD-10 Code Group D)**

| | | Laterality | | |
|---|---|---|---|---|
| **Code** | **Diagnosis** | **Left** | **Right** | **Unspecified** |
| **S40** | **Superficial injury of shoulder and upper arm** | S40.012*,<br>S40.022*,<br>S40.212*,<br>S40.222*,<br>S40.242*,<br>S40.252*,<br>S40.262*,<br>S40.272*,<br>S40.812*,<br>S40.822*,<br>S40.842*,<br>S40.852*,<br>S40.862*,<br>S40.872*,<br>S40.912*,<br>S40.922* | S40.011*,<br>S40.021*,<br>S40.211*,<br>S40.221*,<br>S40.241*,<br>S40.251*,<br>S40.261*,<br>S40.271*,<br>S40.811*,<br>S40.821*,<br>S40.841*,<br>S40.851*,<br>S40.861*,<br>S40.871*,<br>S40.911*,<br>S40.921* | S40.019*,<br>S40.029*,<br>S40.219*,<br>S40.229*,<br>S40.249*,<br>S40.259*,<br>S40.269*,<br>S40.279*,<br>S40.819*,<br>S40.829*,<br>S40.849*,<br>S40.859*,<br>S40.869*,<br>S40.879*,<br>S40.919*,<br>S40.929* |
| **S41** | **Open wound of shoulder and upper arm** | S41.002*,<br>S41.012*,<br>S41.022*,<br>S41.032*,<br>S41.042*,<br>S41.052*,<br>S41.102*,<br>S41.112*,<br>S41.122*,<br>S41.132*,<br>S41.142*,<br>S41.152* | S41.001*,<br>S41.011*,<br>S41.021*,<br>S41.031*,<br>S41.041*,<br>S41.051*,<br>S41.101*,<br>S41.111*,<br>S41.121*,<br>S41.131*,<br>S41.141*,<br>S41.151* | S41.009*,<br>S41.019*,<br>S41.029*,<br>S41.039*,<br>S41.049*,<br>S41.059*,<br>S41.109*,<br>S41.119*,<br>S41.129*,<br>S41.139*,<br>S41.149*,<br>S41.159* |
| **S42** | **Fracture of shoulder and upper arm** | | | |
| **S42.0** | **Fracture of clavicle** | S42.002*,<br>S42.012*,<br>S42.022*,<br>S42.032*,<br>S42.015*,<br>S42.015*,<br>S42.018*,<br>S42.018*,<br>S42.034*,<br>S42.035* | S42.001*,<br>S42.011*,<br>S42.021*,<br>S42.031*,<br>S42.014*,<br>S42.014*,<br>S42.017*,<br>S42.017*,<br>S42.024*,<br>S42.034* | S42.009*,<br>S42.013*,<br>S42.016*,<br>S42.019*,<br>S42.023*,<br>S42.029*,<br>S42.026*,<br>S42.039*,<br>S42.033*,<br>S42.036* |
| **S42.1** | **Fracture of scapula** | S42.102*,<br>S42.112*,<br>S42.115*,<br>S42.122*,<br>S42.125*, | S42.101*,<br>S42.111*,<br>S42.114*,<br>S42.121*,<br>S42.124*, | S42.109*,<br>S42.113*,<br>S42.116*,<br>S42.123*,<br>S42.126*, |

| | | | | |
|---|---|---|---|---|
| | | S42.132*, | S42.131*, | S42.133*, |
| | | S42.135*, | S42.134*, | S42.136*, |
| | | S42.142*, | S42.141*, | S42.143*, |
| | | S42.145*, | S42.144*, | S42.146*, |
| | | S42.152*, | S42.151*, | S42.153*, |
| | | S42.155*, | S42.154*, | S42.156*, |
| | | S42.192* | S42.191* | S42.199* |
| **S42.2** | **Fracture of upper end of humerus** | S42.202*, | S42.201*, | S42.209*, |
| | | S42.212*, | S42.211*, | S42.213*, |
| | | S42.215*, | S42.214*, | S42.216*, |
| | | S42.222*, | S42.221*, | S42.223*, |
| | | S42.225*, | S42.224*, | S42.226*, |
| | | S42.232*, | S42.231*, | S42.239*, |
| | | S42.242*, | S42.241*, | S42.249*, |
| | | S42.252*, | S42.251*, | S42.253*, |
| | | S42.255*, | S42.254*, | S42.256*, |
| | | S42.262*, | S42.261*, | S42.263*, |
| | | S42.265*, | S42.264*, | S42.266*, |
| | | S42.272*, | S42.271*, | S42.279*, |
| | | S42.292*, | S42.291*, | S42.293*, |
| | | S42.295* | S42.294* | S42.296* |
| **S42.3** | **Fracture of shaft of humerus** | S42.302*, | S42.301*, | S42.309*, |
| | | S42.312*, | S42.311*, | S42.319*, |
| | | S42.322*, | S42.321*, | S42.323*, |
| | | S42.325*, | S42.324*, | S42.326*, |
| | | S42.332*, | S42.331*, | S42.333*, |
| | | S42.335*, | S42.334*, | S42.336*, |
| | | S42.342*, | S42.341*, | S42.343*, |
| | | S42.345*, | S42.344*, | S42.346*, |
| | | S42.352*, | S42.351*, | S42.353*, |
| | | S42.355*, | S42.354*, | S42.356*, |
| | | S42.362*, | S42.361*, | S42.363*, |
| | | S42.365*, | S42.364*, | S42.366*, |
| | | S42.392* | S42.391* | S42.399* |
| **S42.4** | **Fracture of lower end of humerus** | S42.402*, | S42.401*, | S43.409*, |
| | | S42.412*, | S42.411*, | S43.413*, |
| | | S42.415*, | S42.414*, | S43.416*, |
| | | S42.422*, | S42.421*, | S43.423*, |
| | | S42.425*, | S42.424*, | S43.426*, |
| | | S42.432*, | S42.431*, | S43.433*, |
| | | S42.435*, | S42.434*, | S43.436*, |
| | | S42.442*, | S42.441*, | S43.443*, |
| | | S42.445*, | S42.444*, | S43.446*, |
| | | S42.448*, | S42.447*, | S43.449*, |
| | | S42.452*, | S42.451*, | S43.453*, |
| | | S42.455*, | S42.454*, | S43.456*, |
| | | S42.462*, | S42.461*, | S43.463*, |
| | | S42.465*, | S42.464*, | S43.466*, |
| | | S42.472*, | S42.471*, | S43.473*, |
| | | S42.475*, | S42.474*, | S43.476*, |
| | | S42.482*, | S42.481*, | S43.489*, |
| | | S42.492*, | S42.491*, | S43.493*, |
| | | S42.495* | S42.494* | S43.496* |

| | | | | |
|---|---|---|---|---|
| **S42.9** | **Fracture of shoulder girdle part unspecified** | S42.92X* | S42.91X* | S42.90X* |
| **S43** | **Dislocation and sprain of joints and ligaments of shoulder girdle** | S43.002*,<br>S43.005*,<br>S43.012*,<br>S43.015*,<br>S43.022*,<br>S43.025*,<br>S43.032*,<br>S43.035*,<br>S43.082*,<br>S43.085*,<br>S43.102*,<br>S43.112*,<br>S43.122*,<br>S43.132*,<br>S43.152*,<br>S43.202*,<br>S43.205*,<br>S43.302*,<br>S43.305*,<br>S43.312*,<br>S43.315*,<br>S43.392*,<br>S43.395*,<br>S43.402*,<br>S43.412*,<br>S43.422*,<br>S43.432*,<br>S43.492*,<br>S43.52X*,<br>S43.62X*,<br>S43.82X*,<br>S43.92X* | S43.001*,<br>S43.004*,<br>S43.011*,<br>S43.014*,<br>S43.021*,<br>S43.024*,<br>S43.031*,<br>S43.034*,<br>S43.081*,<br>S43.084*,<br>S43.101*,<br>S43.111*,<br>S43.121*,<br>S43.131*,<br>S43.151*,<br>S43.201*,<br>S43.204*,<br>S43.301*,<br>S43.304*,<br>S43.311*,<br>S43.314*,<br>S43.391*,<br>S43.394*,<br>S43.401*,<br>S43.411*,<br>S43.421*,<br>S43.431*,<br>S43.491*,<br>S43.51X*,<br>S43.61X*,<br>S43.81X*,<br>S43.91X* | S43.003*,<br>S43.006*,<br>S43.013*,<br>S43.016*,<br>S43.023*,<br>S43.026*,<br>S43.033*,<br>S43.036*,<br>S43.083*,<br>S43.086*,<br>S43.109*,<br>S43.119*,<br>S43.129*,<br>S43.139*,<br>S43.159*,<br>S43.203*,<br>S43.206*,<br>S43.303*,<br>S43.306*,<br>S43.313*,<br>S43.316*,<br>S43.393*,<br>S43.396*,<br>S43.409*,<br>S43.419*,<br>S43.429*,<br>S43.439*,<br>S43.499*,<br>S43.50X*,<br>S43.60X*,<br>S43.80X*,<br>S43.90X* |
| **S44** | **Injury of nerves at shoulder and upper arm level** | S44.32X*,<br>S44.8X2*,<br>S44.92X* | S44.31X*,<br>S44.8X1*,<br>S44.91X* | S44.30X*,<br>S44.8X9*,<br>S44.90X* |
| **S45** | **Injury of blood vessels at shoulder and upper arm level** | S45.002*,<br>S45.012*,<br>S45.092*,<br>S45.202*,<br>S45.212*,<br>S45.292*,<br>S45.802*,<br>S45.812*,<br>S45.892*,<br>S45.902*,<br>S45.912*,<br>S45.992* | S45.001*,<br>S45.011*,<br>S45.091*,<br>S45.201*,<br>S45.211*,<br>S45.291*,<br>S45.801*,<br>S45.811*,<br>S45.891*,<br>S45.901*,<br>S45.911*,<br>S45.991* | S45.009*,<br>S45.019*,<br>S45.099*,<br>S45.209*,<br>S45.219*,<br>S45.299*,<br>S45.809*,<br>S45.819*,<br>S45.899*,<br>S45.909*,<br>S45.919*,<br>S45.999* |
| **S46** | **Injury of muscle fascia and tendon at shoulder and upper arm level** | S46.002*,<br>S46.012*,<br>S46.022*, | S46.001*,<br>S46.011*,<br>S46.021* | S46.009*,<br>S46.019*,<br>S46.029* |

| | | | | |
|---|---|---|---|---|
| | | S46.092*, S46.802*, S46.812*, S46.822*, S46.892*, S46.902*, S46.912*, S46.922*, S46.982*, S46.992* | S46.091*, S46.801*, S46.811*, S46.821*, S46.891*, S46.901*, S46.911*, S46.921*, S46.981*, S46.991* | S46.099*, S46.809*, S46.819*, S46.829*, S46.899*, S46.909*, S46.919*, S46.929*, S46.989*, S46.999* |
| **S47** | **Crushing injury of shoulder and upper arm** | S47.2XX* | S47.1XX* | S47.9XX* |
| **S48** | **Traumatic amputation of shoulder and upper arm** | S48.012*, S48.022*, S48.112*, S48.122*, S48.912*, S48.922* | S48.011*, S48.021*, S48.111*, S48.121*, S48.911*, S48.921* | S48.019*, S48.029*, S48.119*, S48.129*, S48.919*, S48.929* |
| **S49** | **Other and unspecified injuries of shoulder and upper arm** | S49.002*, S49.012*, S49.022*, S49.032*, S49.042*, S49.092*, S49.102*, S49.112*, S49.122*, S49.132*, S49.142*, S49.192*, S49.82X*, S49.92X* | S49.001*, S49.011*, S49.021*, S49.031*, S49.041*, S49.091*, S49.101*, S49.111*, S49.121*, S49.131*, S49.141*, S49.191*, S49.81X*, S49.91X* | S49.009*, S49.019*, S49.029*, S49.039*, S49.049*, S49.099*, S49.109*, S49.119*, S49.129*, S49.139*, S49.149*, S49.199*, S49.80X*, S49.90X* |

### Multimedia Appendix 2:

#### SIRVA abstraction form

1. SCK Study ID (pre-populated) ______________________

2. VSD Study ID (pre-populated) ______________________

##### A. BACKGROUND INFORMATION

3. Abstractor initials ______________________

4. Abstraction date ______________________

##### B. VACCINATIONWe are interested in vaccines administered in 2016-2017. The vaccine types and date are prepopulated.

5-1). Vaccine type (pre-populated) ______________________

5-2). ______________________

5-3). ______________________

5-4). ______________________

6. Vaccination date (pre-populated) ______________________

7. Was there a vaccine given on this day?
- ◯ Yes
- ◯ No
- ◯ Unknown

8. Are there clinic notes available for the vaccination visit?
- ◯ Yes
- ◯ No

(If there is registry information but no clinic notes, select "No")

8-1). If yes, was vaccination documented in the chart notes? (e.g., nursing notes, etc.)
- ◯ Yes
- ◯ Encounter notes available but no mention of vaccination
- ◯ Documented in the immunization record only
- ◯ Mentioned in clinic notes but not noted as given - documented in the immunization record
- ◯ Other

| | |
|---|---|
| 8-2). Please copy and paste the relevant statement for the vaccine(s) given in the chart notes. | ______________________________ |
| 8-3). Give the statement to specify Other. | ______________________________ |
| 9. Was the route of the administration for the vaccine intramuscular (IM)? | ◯ Yes<br>◯ No<br>◯ Unknown |
| 10. Was the vaccine administered in the deltoid muscle (i.e., the upper arm)? | ◯ Yes<br>◯ No<br>◯ Unknown |
| 11. Vaccination side from SCK data (pre-populated) | ◯ Right<br>◯ Left<br>◯ Both<br>◯ Unknown |
| 12. Vaccination side noted in the chart/registry | ◯ Right<br>◯ Left<br>◯ Both<br>◯ Unknown |
| 13. Shoulder of interest<br><br>If the side from the chart/registry (right, left, or both) does not agree with the prepopulated value above, then use the chart value. If no chart value is available, then use the prepopulated value. Hereafter, this will be referred to as the "shoulder of interest" and will be populated in questions about the side of symptoms/injury. | ◯ Right<br>◯ Left<br>◯ Both<br>◯ Unknown |
| 14. The setting of vaccination | ◯ Clinic/Doctor's Office<br>◯ Urgent Care<br>◯ Emergency Room<br>◯ Hospital Inpatient<br>◯ Pharmacy<br>◯ Work<br>◯ Flu vaccine clinic/booth<br>◯ Other (specify)<br>◯ Unknown<br>(Select the location that best describes where the patient was vaccinated. If the patient is a healthcare worker who received his/her vaccination at the place of employment, select "Work.") |
| 14-1). Specify other location of vaccination. | ________________________ |

| | |
|---|---|
| 15. Credentials of the vaccinator | ◯ LVN (Licensed Vocational Nurse)/LPN (Licensed Practical Nurse)<br>◯ MA (Medical Assistant)<br>◯ MD/DO (Physician)<br>◯ NP (Nurse Practitioner)<br>◯ PA (Physician's Assistant)<br>◯ Pharmacist<br>◯ Pharmacy Assistant<br>◯ RN (Registered Nurse)<br>◯ Unknown<br>◯ Other (specify) |
| 15-1). Specify other credentials of the vaccinator. | ____________________________ |
| 14R. The setting of vaccination in RIGHT shoulder/arm | ◯ Clinic/Doctor's Office<br>◯ Urgent Care<br>◯ Emergency Room<br>◯ Hospital Inpatient<br>◯ Pharmacy<br>◯ Work<br>◯ Flu vaccine clinic/booth<br>◯ Other (specify)<br>◯ Unknown<br>(Select the location that best describes where the patient was vaccinated. If the patient is a healthcare worker who received his/her vaccination at the place of employment, select "Work.") |
| 14R-1). Specify other locations of vaccination in RIGHT shoulder/arm. | ____________________________ |
| 15R. Credentials of the vaccinator in RIGHT shoulder/arm | ◯ LVN (Licensed Vocational Nurse)/LPN (Licensed Practical Nurse)<br>◯ MA (Medical Assistant)<br>◯ MD/DO (Physician)<br>◯ NP (Nurse Practitioner)<br>◯ PA (Physician's Assistant)<br>◯ Pharmacist<br>◯ Pharmacy Assistant<br>◯ RN (Registered Nurse)<br>◯ Unknown<br>◯ Other (specify) |
| 15R-1). Specify other credentials of vaccinator in RIGHT shoulder/arm. | ____________________________ |

| | |
|---|---|
| 14L. The setting of vaccination in LEFT shoulder/arm | ◯ Clinic/Doctor's Office<br>◯ Urgent Care<br>◯ Emergency Room<br>◯ Hospital Inpatient<br>◯ Pharmacy<br>◯ Work<br>◯ Flu vaccine clinic/booth<br>◯ Other (specify)<br>◯ Unknown<br>(Select the location that best describes where the patient was vaccinated. If the patient is a healthcare worker who received his/her vaccination at the place of employment, select "Work.") |
| 14L-1). Specify other locations of vaccination in LEFT shoulder/arm. | ______________________ |
| 15L. Credentials of the vaccinator in LEFT shoulder/arm | ◯ LVN (Licensed Vocational Nurse)/LPN (Licensed Practical Nurse)<br>◯ MA (Medical Assistant)<br>◯ MD/DO (Physician)<br>◯ NP (Nurse Practitioner)<br>◯ PA (Physician's Assistant)<br>◯ Pharmacist<br>◯ Pharmacy Assistant<br>◯ RN (Registered Nurse)<br>◯ Unknown<br>◯ Other (specify) |
| 15L-1). Specify other credentials of vaccinator in LEFT shoulder/arm. | ______________________ |
| 16. Were any problems with the vaccine injection noted (e.g., administration error)? | ◯ Yes<br>◯ No |
| 16-1). If yes, specify errors noted. | ______________________ |
| 17. Did the patient have any post-vaccination shoulder or arm (same as vaccinated) complaints noted at this visit? | ◯ Yes<br>◯ No |
| 18. Does the note mention any previous shoulder symptoms? | ◯ Yes<br>◯ No |
| 18-1). Side of previous shoulder symptoms | ◯ Right<br>◯ Left<br>◯ Both<br>◯ Unknown |
| 18-2). Please copy and paste the relevant statement for previous shoulder symptoms and side mentioned from the chart notes. | ______________________ |

#### C. MEDICAL ENCOUNTERS ON THE SAME DAY AS VACCINATION

| | |
|---|---|
| 19. Were there any other provider visits on the same calendar day as vaccination?<br><br>Please only include encounters with patient/provider interaction, providers can include physicians, nurses, physical therapists, and other allied health workers. | ◯ Yes<br>◯ No<br>◯ Unknown |
| 20. Type of encounter | ◯ Outpatient (primary care and specialty care office visit)<br>◯ Urgent care<br>◯ Emergency room<br>◯ Hospital inpatient<br>◯ Phone call<br>◯ Email<br>◯ Other (specify) |
| 20-1). Specify other. | ______________________________ |
| 21. Provider type | ◯ Primary Care<br>◯ ER/Urgent Care/Inpatient<br>◯ Shoulder specialist<br>◯ Non-shoulder specialist<br>◯ Other (specify) |
| 21-1). Specify other. | ______________________________ |
| 22. Were any shoulder symptoms noted at this visit? | ◯ Yes<br>◯ No |
| 22-1). On which side were the shoulder symptoms noted? | ◯ Right<br>◯ Left<br>◯ Both<br>◯ Unknown |
| 23. List any shoulder or arm diagnoses made at this visit. (Leave this blank if no diagnosis is given). | ______________________________ |
| 24. When did the shoulder or arm symptoms start?<br><br>Please be as specific as possible. If a date is leave given, give the date.<br>- If a duration is given (e.g., "two weeks ago"), count backward that amount of time from the visit date and use that as the start date, even if the duration given was inexact (e.g., "about two weeks ago").<br>- If a range is given (e.g., "one to two weeks ago"), give the earliest possible start date (in that case, two weeks prior to the appointment).<br>- Use the calendar box to select a date and write the exact statement regarding the start date or symptom duration in the text box. | ______________________________<br>(If the duration of symptoms is not discussed, this blank.) |

| | |
|---|---|
| 25. Is this date exact or an estimation? | ◯ Exact<br>◯ Estimate<br>(Select "estimate" if symptom duration/onset date was not discussed.) |
| 26. Is this symptom onset date before OR after vaccination? | ◯ Before<br>◯ After<br>◯ Same |
| 26-1). If the symptom onset date is the same as the vaccination date, is symptom onset before or after the vaccination? | ◯ Before<br>◯ After |
| 27. Give the statement regarding the description of the symptoms and start date or symptom duration as it is in the chart. If the duration is not discussed, enter that here as well. | ___________________________ |
| 28. Was the shoulder or arm diagnosis attributed to the vaccination by the provider? | ◯ Yes<br>◯ No<br>◯ Not stated |
| 29. Please copy and paste the relevant statement for any cause of the shoulder symptoms from the chart note AND save the full visit note (unredacted) to S: drive as part of a case packet. | ___________________________ |
| 24R. When did the RIGHT shoulder or arm symptoms start?<br><br>Please be as specific as possible. If a date is given, give the date.<br>- If a duration is given (e.g., "two weeks ago"), count backward that amount of time from the visit date and use that as the start date, even if the duration given was inexact (e.g., "about two weeks ago").<br>- If a range is given (e.g., "one to two weeks ago"), give the earliest possible start date (in that case, two weeks prior to the appointment).<br>- Use the calendar box to select a date and write the exact statement regarding the start date or symptom duration in the text box. | ______________________<br>(If the duration of symptoms is not discussed, leave this blank.) |
| 25R. Is this date exact or an estimation? | ◯ Exact<br>◯ Estimate<br>(Select "estimate" if symptom duration/onset date was not discussed.) |
| 26R. Is the RIGHT shoulder symptom onset date before OR after vaccination? | ◯ Before<br>◯ After<br>◯ Same |
| 26R-1). If the RIGHT shoulder symptom onset date is the same as the vaccination date, is symptom onset before or after the vaccination? | ◯ Before<br>◯ After |

| | |
|---|---|
| 27R. Give the statement regarding the description of the RIGHT shoulder symptoms and start date or RIGHT shoulder symptom duration as it is in the chart. If the duration is not discussed, enter that here as well. | ________________________________ |
| 28R. Was the RIGHT shoulder or arm diagnosis attributed to the vaccination by the provider? | ◯ Yes<br>◯ No<br>◯ Not stated |
| 29R. Please copy and paste the relevant statement for any cause of the RIGHT shoulder symptoms from the chart note AND save the full visit note (unredacted) to S: drive as part of a case packet. | ________________________________ |
| 24L. When did the LEFT shoulder or arm symptoms start?<br><br>Please be as specific as possible. If a date is given, give the date.<br>- If a duration is given (e.g., "two weeks ago"), count backward that amount of time from the visit date and use that as the start date, even if the duration given was inexact (e.g., "about two weeks ago").<br>- If a range is given (e.g., "one to two weeks ago"), gives the earliest possible start date (in that case, two weeks prior to the appointment).<br>- Use the calendar box to select a date and write the exact statement regarding the start date or symptom duration in the text box. | _________________________<br>(If the duration of symptoms is not discussed, leave this blank.) |
| 25L. Is this date exact or an estimation? | ◯ Exact<br>◯ Estimate<br>(Select "estimate" if symptom duration/onset date was not discussed.) |
| 26L. Is the LEFT shoulder symptom onset date before OR after vaccination? | ◯ Before<br>◯ After<br>◯ Same |
| 26L-1). If the LEFT shoulder symptom onset date is the same as the vaccination date, is LEFT shoulder symptom onset before or after the vaccination? | ◯ Before<br>◯ After |
| 27L. Give the statement regarding the description of the LEFT shoulder symptoms and start date or LEFT shoulder symptom duration as it is in the chart. If the duration is not discussed, enter that here as well. | ________________________________ |
| 28L. Was the LEFT shoulder or arm diagnosis attributed to the vaccination by the provider? | ◯ Yes<br>◯ No<br>◯ Not stated |
| 29L. Please copy and paste the relevant statement for any cause of the LEFT shoulder symptoms from the chart note AND save the full visit note (unredacted) to S: drive as part of a case packet. | ________________________________ |

#### D. PRE-EXISTING SHOULDER CONDITIONS DOCUMENTED PRIOR TO VACCINATION DATE

**Working backward from the date of vaccination to 6 months prior to vaccination, look for any visits for shoulder or upper arm problems, symptoms, or injuries. Do not worry about any injuries or symptoms to the elbow, forearm, wrist, or hand.**

| | |
|---|---|
| 30. Any prior shoulder/upper arm symptoms found? | ◯ Yes<br>◯ No |
| 30-1). On which side were the shoulder symptoms noted? | ◯ Right<br>◯ Left<br>◯ Both<br>◯ Unknown |
| 31. Visit date of most recent visit for shoulder/upper arm symptoms prior to vaccination | ______________________ |
| 32. Provider type | ◯ Primary Care<br>◯ ER/Urgent Care/Inpatient<br>◯ Shoulder specialist<br>◯ Non-shoulder specialist<br>◯ Other (specify) |
| 32-1). Specify other providers. | ______________________ |
| 33. Was there a previous diagnosis of the following shoulder injuries in the above shoulder in 6 months prior to vaccination?<br><br>- Adhesive capsulitis/ frozen shoulder<br>- Bone erosion<br>- Bursitis<br>- Humerus fractures<br>- Impingement<br>- Left shoulder joint pain<br>- Osteitis<br>- Osteolysis<br>- Osteonecrosis<br>- Periosteal reactions<br>- Pseudoseptic arthritis<br>- Right shoulder joint pain<br>- Rotator cuff syndrome<br>- Shoulder joint effusion<br>- Synovitis/ tenosynovitis<br>- Tendinitis/ tendinosis/ tendonitis/ tendinopathy<br>- Torn rotator cuff | ◯ Yes<br>◯ No |
| 34. Was there a previous shoulder/upper arm diagnosis other than the above shoulder injuries (Q33) in the above shoulder? | ◯ Yes<br>◯ No |
| 34-1). Specify other shoulder/upper arm diagnoses (list all shoulder/upper arm diagnoses given during the 6 months prior to vaccination). | ______________________ |
| 31R. Visit date of most recent visit for RIGHT shoulder/upper arm symptoms prior to vaccination | ______________________ |

| | |
|---|---|
| 32R. Provider type | ◯ Primary Care<br>◯ ER/Urgent Care/Inpatient<br>◯ Shoulder specialist<br>◯ Non-shoulder specialist<br>◯ Other (specify) |
| 32R-1). Specify other provider. | ______________________________ |
| 33R. Was there a previous diagnosis of the following shoulder injuries in the RIGHT shoulder in 6 months prior to vaccination?<br><br>- Adhesive capsulitis/ frozen shoulder<br>- Bone erosion<br>- Bursitis<br>- Humerus fractures<br>- Impingement<br>- Left shoulder joint pain<br>- Osteitis<br>- Osteolysis<br>- Osteonecrosis<br>- Periosteal reactions<br>- Pseudoseptic arthritis<br>- Right shoulder joint pain<br>- Rotator cuff syndrome<br>- Shoulder joint effusion<br>- Synovitis/ tenosynovitis<br>- Tendinitis/ tendinosis/ tendonitis/ tendinopathy<br>- Torn rotator cuff | ◯ Yes<br>◯ No |
| 34R. Was there a previous shoulder/upper arm diagnosis other than the above shoulder injuries (Q33R) in the RIGHT shoulder? | ◯ Yes<br>◯ No |
| 34R-1). Specify other RIGHT shoulder/upper arm diagnoses (list all shoulder/upper arm diagnoses given during the 6 months prior to vaccination). | ______________________________ |
| 31L. Visit date of most recent visit for LEFT shoulder/upper arm symptoms prior to vaccination | ______________________________ |
| 32L. Provider type | ◯ Primary Care<br>◯ ER/Urgent Care/Inpatient<br>◯ Shoulder specialist<br>◯ Non-shoulder specialist<br>◯ Other (specify) |
| 32L-1). Specify other providers. | ______________________________ |

| | |
|---|---|
| 33L. Was there a previous diagnosis of the following shoulder injuries in the LEFT shoulder in 6 months prior to vaccination?<br><br>- Adhesive capsulitis/ frozen shoulder<br>- Bone erosion<br>- Bursitis<br>- Humerus fractures<br>- Impingement<br>- Left shoulder joint pain<br>- Osteitis<br>- Osteolysis<br>- Osteonecrosis<br>- Periosteal reactions<br>- Pseudoseptic arthritis<br>- Right shoulder joint pain<br>- Rotator cuff syndrome<br>- Shoulder joint effusion<br>- Synovitis/ tenosynovitis<br>- Tendinitis/ tendinosis/ tendonitis/ tendinopathy<br>- Torn rotator cuff | ◯ Yes<br>◯ No |
| 34L. Was there a previous LEFT shoulder/upper arm diagnosis other than the above shoulder injuries (Q33L) in the above shoulder? | ◯ Yes<br>◯ No |
| 34L-1). Specify other LEFT shoulder/upper arm diagnoses (list all shoulder/upper arm diagnoses given during the 6 months prior to vaccination). | ______________________________ |

#### E. NEXT MEDICAL ENCOUNTER AFTER VACCINATION

**We are interested in the first medical encounter that happened on a date other than the vaccination date, within 30 days of vaccination, even if it is not related to the shoulder/arm or vaccination.**

| | |
|---|---|
| 35. Is there an encounter within 30 days of vaccination? | ◯ Yes<br>◯ No |
| 36. Date of next medical encounter | ______________________________ |
| 37. Type of encounter | ◯ Outpatient (primary care and specialty care office visit)<br>◯ Urgent care<br>◯ Emergency room<br>◯ Hospital inpatient<br>◯ Phone call<br>◯ Email<br>◯ Other (specify) |
| 37-1). Specify other. | ______________________________ |

| | |
|---|---|
| 38. Provider type | ◯ Primary Care<br>◯ ER/Urgent Care/Inpatient<br>◯ Shoulder specialist<br>◯ Non-shoulder specialist<br>◯ Other (specify) |
| 38-1). If other, specify. | ____________________________ |
| 39. What was the chief complaint of this visit? | ____________________________ |
| 40. Were any shoulder/upper arm symptoms documented as part of this encounter? | ◯ Yes<br>◯ No |
| 40-1). On which side did the patient complain of shoulder/upper arm symptoms? | ◯ Right<br>◯ Left<br>◯ Both<br>◯ Unknown |

#### F. FIRST SHOULDER/UPPER ARM VISIT WITHIN 30 DAYS POST-VACCINATION

**Look for the first clinic visit for shoulder/upper arm symptoms, within 30 days of vaccination. If the visit is clearly NOT for the shoulder of interest, go to the first visit that is for the shoulder of interest/both shoulders/laterality is unclear.**

**If the answer to the previous question (Q40. Did the patient complain of shoulder/upper arm symptoms as part of this encounter?) is Yes, answer these questions based on that visit. If the answer was No, look for the next clinic visit that involves shoulder/upper arm symptoms and answer the questions based on that visit.**

| | |
|---|---|
| 41. Is there a shoulder/upper arm visit within 30 days of the vaccination date? | Yes<br>No |
| 42. Date of the first shoulder/upper arm visit. | ____________________________<br>(If there are no shoulder visits please enter '09/09/9999'.) |
| 43. Type of encounter | ◯ Outpatient (primary care and specialty care office visit)<br>◯ Urgent care<br>◯ Emergency room<br>◯ Hospital inpatient<br>◯ Other (specify) |
| 43-1). Specify other. | ____________________________ |

| | |
|---|---|
| 44. Type of provider | ◯ Orthopedics<br>◯ Rheumatologist<br>◯ Physical Medicine & Rehabilitation<br>◯ Sports Medicine<br>◯ Primary Care Physician (eg, internal medicine, family medicine, pediatrics)<br>◯ Physical Therapy<br>◯ Occupational Therapy<br>◯ Other (specify) |
| 44-1). Specify other provider type. | ____________________________ |
| 45. Which side has the symptoms? | ◯ Right<br>◯ Left<br>◯ Both<br>◯ Unknown |
| 46. When did the shoulder or arm symptoms start?<br><br>Please be as specific as possible. If a date is given, give the date. If a duration is given (e.g., "two weeks ago") count backwards that amount of time from the visit date and use that as the start date, even if the duration given was inexact (e.g., "about two weeks ago"). If a range is given (e.g., "one to two weeks ago") give the earliest possible start date (in that case, two weeks prior to the appointment). Use the calendar box to select a date and write the exact statement regarding the start date or symptom duration in the text box. | ____________________________<br>(Leave blank if symptom duration or onset date was not discussed at this visit.) |
| 47. Is this date exact or an estimation? | ◯ Exact<br>◯ Estimate<br>(Select "estimate" if the duration of symptoms was not discussed.) |
| 48. Is this pre-existing condition? | ◯ Yes<br>◯ No<br>◯ Possible<br>◯ Unknown |
| 49. Were the pre-existing shoulder/arm symptoms noted to recur or worsen following vaccination?<br><br>If no, please STOP here and END abstraction. | ◯ Yes<br>◯ No<br>◯ Possible<br>◯ Unknown |
| 50. Did the recurrence or worsening of symptoms occur within 30 days of vaccination?<br><br>If no, please STOP here and END abstraction. | ◯ Yes<br>◯ No<br>◯ Unknown |
| 51. Give the statement regarding the description of the symptoms, start date or symptom duration as it is in the chart. If duration is not discussed, enter that here as well.<br><br>Please save the full visit note (unredacted) to S: drive as part of a case packet. | ____________________________________ |

| | |
|---|---|
| 52. Check all the symptoms that were reported | ☐ Atrophy<br>☐ Impingement<br>☐ Numbness<br>☐ Pain/Soreness<br>☐ Radiating pain (from a non-shoulder location)<br>☐ Reduced range of motion (ROM)<br>☐ Stiffness<br>☐ Swelling<br>☐ Tingling<br>☐ Weakness<br>☐ Other shoulder symptoms (specify) |
| 52-1). Specify other. | ________________________ |
| 53. Was a specific injury or action, other than vaccination, mentioned as the cause of the shoulder/upper arm symptoms? | ○ Yes<br>○ No |
| 54. Were the shoulder/upper arm symptoms attributed to the vaccine by the patient or provider? | ○ Yes<br>○ No |
| 55. Please copy and paste the relevant statement from the chart note for any cause of the symptoms AND save the full visit note (unredacted) to S: drive as part of a case packet. | ______________________________ |
| 56. Was vaccination mentioned at all in the visit note? | ○ Yes<br>○ No |
| 57. Was the vaccine type specified in the statement? | ○ Yes<br>○ No |
| 57-1). If the above question is "Yes", select the vaccine type from the following list. | ☐ Influenza<br>☐ TDAP<br>☐ Pneumococcal<br>☐ Hepatitis A or B<br>☐ HPV<br>☐ Meningococcal<br>☐ Other (specify) |
| 57-1a). Specify other. | ________________________ |
| 57-2). Does the vaccine type state match to the one listed in Q5? | ○ Yes<br>○ No |
| 58. Were any symptoms documented described as being related to a vaccine injection? Select as many as apply. | ☐ Induration/Hard mass<br>☐ Erythema/Swelling/Redness<br>☐ Pain/Soreness<br>☐ Rash<br>☐ Other (specify) |
| 58-1). Specify other. | ________________________ |

| | |
|---|---|
| 59. Specify positive/abnormal physical exam findings pertaining to the above shoulder/upper arm. | ______________________________ |
| 60. Was there a diagnosis of the following shoulder injuries in the above shoulder at this visit? Please select as many diagnoses given at this visit. | ☐ Adhesive capsulitis/ frozen shoulder<br>☐ Bone erosion<br>☐ Bursitis<br>☐ Impingement<br>☐ Osteitis<br>☐ Osteolysis<br>☐ Osteonecrosis<br>☐ Periosteal reactions<br>☐ Pseudoseptic arthritis<br>☐ Shoulder joint effusion<br>☐ Synovitis/ tenosynovitis<br>☐ Tendinitis/ tendinosis/ tendonitis/ tendinopathy<br>☐ Torn rotator cuff<br>☐ Rotator cuff syndrome<br>☐ Right shoulder joint pain<br>☐ Left shoulder joint pain<br>☐ Humerus fractures<br>☐ Other<br>☐ No shoulder injury diagnosis |
| 60-1). Specify other shoulder/upper arm diagnoses (If there is no Other diagnosis, please leave blank.) | ________________________ |
| 46R. When did the RIGHT shoulder or arm symptoms start?<br><br>Please be as specific as possible. If a date is given, give the date. If a duration is given (e.g., "two weeks ago") count backward that amount of time from the visit date and use that as the start date, even if the duration given was inexact (e.g., "about two weeks ago"). If a range is given (e.g., "one to two weeks ago") give the earliest possible start date (in that case, two weeks prior to the appointment). Use the calendar box to select a date and write the exact statement regarding the start date or symptom duration in the text box. | ________________________<br>(Leave blank if symptom duration or onset date was not discussed at this visit.) |
| 47R. Is this date exact or an estimation? | ◯ Exact<br>◯ Estimate<br>(Select "estimate" if the duration of symptoms was not discussed.) |
| 48R. Is this pre-existing condition? | ◯ Yes<br>◯ No<br>◯ Possible<br>◯ Unknown |
| 49R. Were the pre-existing RIGHT shoulder/arm symptoms noted to recur or worsen following vaccination?<br><br>If no, please STOP here and END abstraction. | ◯ Yes<br>◯ No<br>◯ Possible<br>◯ Unknown |

| | |
|---|---|
| 50R. Did the recurrence or worsening of RIGHT shoulder symptoms occur within 30 days of vaccination?<br><br>If no, please STOP here and END abstraction. | ◯ Yes<br>◯ No<br>◯ Unknown |
| 51R. Give the statement regarding the description of the RIGHT shoulder symptoms, start date or symptom duration as it is in the chart. If duration is not discussed, enter that here as well.<br><br>Please save the full visit note (unredacted) to S: drive as part of a case packet. | ________________________ |
| 52R. Check all the RIGHT shoulder symptoms that were reported | ☐ Atrophy<br>☐ Impingement<br>☐ Numbness<br>☐ Pain/Soreness<br>☐ Radiating pain (from a non-shoulder location)<br>☐ Reduced range of motion (ROM)<br>☐ Stiffness<br>☐ Swelling<br>☐ Tingling<br>☐ Weakness<br>☐ Other shoulder symptoms (specify) |
| 52R-1). Specify other. | ____________________ |
| 53R. Was a specific injury or action, other than vaccination, mentioned as the cause of the RIGHT shoulder/upper arm symptoms? | ◯ Yes<br>◯ No |
| 54R. Were the RIGHT shoulder/upper arm symptoms attributed to the vaccine by the patient or provider? | ◯ Yes<br>◯ No |
| 55R. Please copy and paste the relevant statement from the chart note for any cause of the RIGHT shoulder symptoms AND save the full visit note (unredacted) to S: drive as part of a case packet. | ________________________ |
| 56R. Was vaccination mentioned at all in the visit note? | ◯ Yes<br>◯ No |
| 57R. Was the vaccine type specified in the statement? | ◯ Yes<br>◯ No |
| 57R-1). If the above question is "Yes", select the vaccine type from the following list. | ☐ Influenza<br>☐ TDAP<br>☐ Pneumococcal<br>☐ Hepatitis A or B<br>☐ HPV<br>☐ Meningococcal<br>☐ Other (specify) |
| 57R-1a). Specify other. | ____________________ |

| | |
|---|---|
| 57R-2). Does the vaccine type stated match to the one listed in Q5? | ◯ Yes<br>◯ No |
| 58R. Were any RIGHT shoulder symptoms documented described as being related to a vaccine injection? Select as many as apply. | ☐ Induration/Hard mass<br>☐ Erythema/Swelling/Redness<br>☐ Pain/Soreness<br>☐ Rash<br>☐ Other (specify) |
| 58R-1). Specify other. | ____________________________ |
| 59R. Specify positive/abnormal physical exam findings pertaining to the RIGHT shoulder/upper arm. | ___________________________________ |
| 60R. Was there a diagnosis of the following shoulder injuries in the RIGHT shoulder at this visit? Please select as many diagnoses are given at this visit. | ☐ Adhesive capsulitis/ frozen shoulder<br>☐ Bone erosion<br>☐ Bursitis<br>☐ Impingement<br>☐ Osteitis<br>☐ Osteolysis<br>☐ Osteonecrosis<br>☐ Periosteal reactions<br>☐ Pseudoseptic arthritis<br>☐ Shoulder joint effusion<br>☐ Synovitis/ tenosynovitis<br>☐ Tendinitis/ tendinosis/ tendonitis/ tendinopathy<br>☐ Torn rotator cuff<br>☐ Rotator cuff syndrome<br>☐ Right shoulder joint pain<br>☐ Left shoulder joint pain<br>☐ Humerus fractures<br>☐ Other<br>☐ No shoulder injury diagnosis |
| 60R-1). Specify other RIGHT shoulder/upper arm diagnoses (If there are no Other diagnosis, please leave blank.) | _____________________________ |
| 46L. When did the LEFT shoulder or arm symptoms start?<br><br>Please be as specific as possible. If a date is given, give the date. If a duration is given (e.g., "two weeks ago") count backward that amount of time from the visit date and use that as the start date, even if the duration given was inexact (e.g., "about two weeks ago"). If a range is given (e.g., "one to two weeks ago") give the earliest possible start date (in that case, two weeks prior to the appointment). Use the calendar box to select a date and write the exact statement regarding the start date or symptom duration in the text box. | ____________________________<br>(Leave blank if symptom duration or onset date was not discussed at this visit.) |
| 47L. Is this date exact or an estimation? | ◯ Exact<br>◯ Estimate<br>(Select "estimate" if the duration of symptoms was not discussed.) |

| | |
|---|---|
| 48L. Is this pre-existing condition? | ◯ Yes<br>◯ No<br>◯ Possible<br>◯ Unknown |
| 49L. Were the pre-existing LEFT shoulder/arm symptoms noted to recur or worsen following vaccination?<br><br>If no, please STOP here and END abstraction. | ◯ Yes<br>◯ No<br>◯ Possible<br>◯ Unknown |
| 50L. Did the recurrence or worsening of LEFT shoulder symptoms occur within 30 days of vaccination?<br><br>If no, please STOP here and END abstraction. | ◯ Yes<br>◯ No<br>◯ Unknown |
| 51L. Give the statement regarding the description of the LEFT shoulder symptoms, start date or symptom duration as it is in the chart. If duration is not discussed, enter that here as well.<br><br>Please save the full visit note (unredacted) to S: drive as part of a case packet. | ______________________________ |
| 52L. Check all the LEFT shoulder symptoms that were reported | ☐ Atrophy<br>☐ Impingement<br>☐ Numbness<br>☐ Pain/Soreness<br>☐ Radiating pain (from a non-shoulder location)<br>☐ Reduced range of motion (ROM)<br>☐ Stiffness<br>☐ Swelling<br>☐ Tingling<br>☐ Weakness<br>☐ Other shoulder symptoms (specify) |
| 52L-1). Specify other. | ________________________ |
| 53L. Was a specific injury or action, other than vaccination, mentioned as the cause of the LEFT shoulder/upper arm symptoms? | ◯ Yes<br>◯ No |
| 54L. Were the LEFT shoulder/upper arm symptoms attributed to the vaccine by the patient or provider? | ◯ Yes<br>◯ No |
| 55L. Please copy and paste the relevant statement from the chart note for any cause of the LEFT shoulder symptoms AND save the full visit note (unredacted) to S: drive as part of a case packet. | ______________________________ |
| 56L. Was vaccination mentioned at all in the visit note? | ◯ Yes<br>◯ No |
| 57L. Was the vaccine type specified in the statement? | ◯ Yes<br>◯ No |

| | |
|---|---|
| 57L-1). If the above question is "Yes", select the vaccine type from the following list. | ☐ Influenza<br>☐ TDAP<br>☐ Pneumococcal<br>☐ Hepatitis A or B<br>☐ HPV<br>☐ Meningococcal<br>☐ Other (specify) |
| 57L-1a). Specify other. | ________________________ |
| 57L-2). Does the vaccine type stated match the one listed in Q5? | ◯ Yes<br>◯ No |
| 58L. Were any LEFT shoulder symptoms documented described as being related to a vaccine injection? Select as many as apply. | ☐ Induration/Hard mass<br>☐ Erythema/Swelling/Redness<br>☐ Pain/Soreness<br>☐ Rash<br>☐ Other (specify) |
| 58L-1). Specify other. | ________________________ |
| 59L. Specify positive/abnormal physical exam findings pertaining to the LEFT shoulder/upper arm. | ______________________________ |
| 60L. Was there a diagnosis of the following shoulder injuries in the LEFT shoulder at this visit? Please select as many diagnoses given at this visit. | ☐ Adhesive capsulitis/ frozen shoulder<br>☐ Bone erosion<br>☐ Bursitis<br>☐ Impingement<br>☐ Osteitis<br>☐ Osteolysis<br>☐ Osteonecrosis<br>☐ Periosteal reactions<br>☐ Pseudoseptic arthritis<br>☐ Shoulder joint effusion<br>☐ Synovitis/ tenosynovitis<br>☐ Tendinitis/ tendinosis/ tendonitis/ tendinopathy<br>☐ Torn rotator cuff<br>☐ Rotator cuff syndrome<br>☐ Right shoulder joint pain<br>☐ Left shoulder joint pain<br>☐ Humerus fractures<br>☐ Other<br>☐ No shoulder injury diagnosis |
| 60L-1). Specify other LEFT shoulder/upper arm diagnoses (If there are no Other diagnosis, please leave blank.) | ________________________ |
| 61-1). List all other diagnoses below given at this appointment, even if they don't pertain to the shoulder. | ________________________ |
| 61-2). | ________________________ |

| | |
|---|---|
| 61-3). | ______________________ |
| 61-4). | ______________________ |
| 61-5). | ______________________ |
| 61-6). | ______________________ |
| 61-7). | ______________________ |
| 61-8). | ______________________ |
| 61-9). | ______________________ |
| 61-10). | ______________________ |
| 61-11). Please include all other diagnoses if more than ten. | ____________________________ |

#### G. FIRST SHOULDER/UPPER ARM VISIT WITHIN 31-180 DAYS POST-VACCINATION

**Look for the first clinic visit for shoulder/upper arm symptoms, within 31-180 days of vaccination. If the visit is clearly NOT for the shoulder of interest, go to the first visit that is for the shoulder of interest/both shoulders/laterality is unclear.**

| | |
|---|---|
| 62. Is there a visit related to the shoulder/arm of interest within 31-180 days following vaccination? If no, go to Section H. | ◯ Yes<br>◯ No |
| 63. Date of visit: | ______________________ |
| 64. Type of encounter: | ◯ Outpatient (primary care and specialty care office visit)<br>◯ Urgent care<br>◯ Emergency room<br>◯ Hospital inpatient<br>◯ Other (specify) |
| 64-1). Specify other. | ______________________ |

| | |
|---|---|
| 65. Type of provider: | ◯ Orthopedics<br>◯ Rheumatologist<br>◯ Physical Medicine & Rehabilitation<br>◯ Sports Medicine<br>◯ Primary Care Physician (eg, internal medicine, family medicine, pediatrics)<br>◯ Physical Therapy<br>◯ Occupational Therapy<br>◯ Other (specify) |
| 65-1). Specify other provider types. | ____________________________ |
| 66. Which shoulder was discussed at the encounter? | ◯ Right<br>◯ Left<br>◯ Both<br>◯ Unknown |
| 67. Did the patient note that his/her symptoms had resolved/relieved? | ◯ Resolved<br>◯ Improved<br>◯ No resolution/improvement<br>◯ Other (e.g., some symptoms improved but other symptoms still present)<br>◯ Unknown |
| 68. Symptoms that the patient is still experiencing: | ☐ Atrophy<br>☐ Impingement<br>☐ Numbness<br>☐ Pain/Soreness<br>☐ Radiating pain (from non-shoulder location)<br>☐ Reduced range of motion (ROM)<br>☐ Stiffness<br>☐ Swelling<br>☐ Tingling<br>☐ Weakness<br>☐ Other shoulder symptoms (specify) |
| 68-1). Specify other. | ____________________________ |
| 69. Was a specific injury or action, other than vaccination, mentioned as the cause of the shoulder/upper arm symptoms? | ◯ Yes<br>◯ No |
| 70. Were the shoulder/upper arm symptoms attributed to the vaccine by the patient or provider? | ◯ Yes<br>◯ No |
| 71. Please copy and paste the relevant statement from the chart note for any cause of the symptoms AND save the full visit note (unredacted) to S: drive as part of a case packet. | ___________________________________ |
| 72. Was vaccination mentioned at all in the visit note? | ◯ Yes<br>◯ No |
| 73. Was the vaccine type specified in the statement? | ◯ Yes<br>◯ No |

| | |
|---|---|
| 73-1). If the above question is "Yes", select the vaccine type from the following list. | ☐ Influenza<br>☐ TDAP<br>☐ Pneumococcal<br>☐ Hepatitis A or B<br>☐ HPV<br>☐ Meningococcal<br>☐ Other (specify) |
| 73-1a). Specify other. | __________________________ |
| 73-2). Does the vaccine type state match the one listed in Q5? | ◯ Yes<br>◯ No |
| 67R. Did the patient note that his/her RIGHT shoulder symptoms had resolved/relieved? | ◯ Resolved<br>◯ Improved<br>◯ No resolution/improvement<br>◯ Other (e.g., some symptoms improved but other symptoms still present)<br>◯ Unknown |
| 68R. RIGHT shoulder symptoms that the patient is still experiencing: | ☐ Atrophy<br>☐ Impingement<br>☐ Numbness<br>☐ Pain/Soreness<br>☐ Radiating pain (from non-shoulder location)<br>☐ Reduced range of motion (ROM)<br>☐ Stiffness<br>☐ Swelling<br>☐ Tingling<br>☐ Weakness<br>☐ Other shoulder symptoms (specify) |
| 68R-1). Specify other. | __________________________ |
| 67L. Did the patient note that his/her LEFT shoulder symptoms had resolved/relieved? | ◯ Resolved<br>◯ Improved<br>◯ No resolution/improvement<br>◯ Other (e.g., some symptoms improved but other symptoms still present)<br>◯ Unknown |
| 68L. LEFT shoulder symptoms that the patient is still experiencing: | ☐ Atrophy<br>☐ Impingement<br>☐ Numbness<br>☐ Pain/Soreness<br>☐ Radiating pain (from non-shoulder location)<br>☐ Reduced range of motion (ROM)<br>☐ Stiffness<br>☐ Swelling<br>☐ Tingling<br>☐ Weakness<br>☐ Other shoulder symptoms (specify) |
| 68L-1). Specify other. | __________________________ |
| 74-1). List all diagnoses given at this appointment, even if they don't pertain to the shoulder. | __________________________ |

74-2). ______________________________

74-3). ______________________________

74-4). ______________________________

74-5). ______________________________

74-6). ______________________________

74-7). ______________________________

74-8). ______________________________

74-9). ______________________________

74-10). ______________________________

74-11). Please include all other diagnoses if more than ten. ____________________________________

**H. Case Definition**

**A SIRVA case is a shoulder injury occurring in the same arm in which a vaccine was injected within the first 7 days following vaccination and lasting more than 30 days following vaccination, with vaccination as one of the possible causes of the shoulder injury.**

**Please use all available records in +/- 6 months from the index date to answer the questions in this section.**

75. In your review, which shoulder/arm was identified with symptoms after vaccination?

- ◯ Right
- ◯ Left
- ◯ Both
- ◯ Unknown

| | |
|---|---|
| 75R. Did a true RIGHT shoulder injury occur? (from the following)<br><br>- Adhesive capsulitis/frozen shoulder<br>- Bone erosion<br>- Bursitis<br>- Humerus fractures<br>- Impingement<br>- Left shoulder joint pain<br>- Osteitis<br>- Osteolysis<br>- Osteonecrosis<br>- Periosteal reactions<br>- Pseudoseptic arthritis<br>- Right shoulder joint pain<br>- Rotator cuff syndrome<br>- Shoulder joint effusion<br>- Synovitis/ tenosynovitis<br>- Tendinitis/ tendinosis/ tendonitis/ tendinopathy<br>- Torn rotator cuff<br>- Other | ◯ Shoulder injury diagnosis code AND physician/patient mention of shoulder symptoms<br>◯ Shoulder injury diagnosis code with no mention of shoulder symptoms by physician/patient<br>◯ Shoulder symptoms mentioned by physician/patient but no shoulder injury diagnosis code<br>◯ No shoulder injury diagnosis code or physician/patient mention of shoulder symptoms |
| 76R-1). List the diagnosis codes from Q75R. | ______________________ |
| 76R-2). | ______________________ |
| 76R-3). | ______________________ |
| 76R-4). | ______________________ |
| 76R-5). | ______________________ |
| 76R-6). | ______________________ |
| 76R-7). | ______________________ |
| 76R-8). | ______________________ |
| 76R-9). | ______________________ |
| 76R-10). | ______________________ |
| 77R. Did the shoulder injury occur in the RIGHT arm in which a vaccine was injected? | ◯ Yes<br>◯ No<br>◯ Unknown |

| | |
|---|---|
| 77R-1). Please copy and paste the relevant statement from the chart notes on the RIGHT shoulder injury. | ________________________________ |
| 78R. Did the RIGHT symptoms of shoulder injury begin within the first 7 days following vaccination?<br><br>(Please be sure to use the most complete and reliable information to answer this question when there are discrepancies with more than one source.) | ◯ Yes<br>◯ No<br>◯ No, but increased severity of pre-existing symptoms in the first 30 days<br>◯ Possible<br>◯ Unknown |
| 78R-1). Please copy and paste the relevant statement from the chart notes on the shoulder injury symptom onset. | ________________________________ |
| 79R. Did the RIGHT shoulder symptoms persist more than 30 days from the date of vaccination?<br><br>(Please be sure to use the most complete and accurate information to answer this question when there are discrepancies with more than one source.) | ◯ Yes<br>◯ No<br>◯ Possible<br>◯ Unknown |
| 79R-1). Please copy and paste the relevant statement from the chart notes on the RIGHT shoulder injury symptom duration. | ________________________________ |
| 80R. What was the cause of the RIGHT shoulder injury?<br><br>(Please select as many causes as documented.) | ☐ Vaccine<br>☐ Incident (e.g. fall, auto accident)<br>☐ Exercise (e.g. exercise, sports)<br>☐ Daily activity (e.g. overuse, lifting a heavy item, work-related injury, side sleeping)<br>☐ Other medical conditions (e.g. arthritis, chest pain radiating to the shoulder)<br>☐ Unknown (e.g. no explicit cause, insidious/aggravating factors- 'worse with exercise') |
| 80R-1). Please copy and paste the relevant statement from the chart notes on the cause of RIGHT shoulder injury (for all of the above responses). | ________________________________ |
| 81R. Does it look like a SIRVA case? | ◯ Yes<br>◯ No<br>◯ Possible<br>◯ Unknown |

| | |
|---|---|
| 75L. Did a true LEFT shoulder injury occur? (from the following)<br><br>- Adhesive capsulitis/frozen shoulder<br>- Bone erosion<br>- Bursitis<br>- Humerus fractures<br>- Impingement<br>- Left shoulder joint pain<br>- Osteitis<br>- Osteolysis<br>- Osteonecrosis<br>- Periosteal reactions<br>- Pseudoseptic arthritis<br>- Right shoulder joint pain<br>- Rotator cuff syndrome<br>- Shoulder joint effusion<br>- Synovitis/ tenosynovitis<br>- Tendinitis/ tendinosis/ tendonitis/ tendinopathy<br>- Torn rotator cuff<br>- Other | ◯ Shoulder injury diagnosis code AND physician/patient mention of shoulder symptoms<br>◯ Shoulder injury diagnosis code with no mention of shoulder symptoms by physician/patient<br>◯ Shoulder symptoms mentioned by physician/patient but no shoulder injury diagnosis code<br>◯ No shoulder injury diagnosis code or physician/patient mention of shoulder symptoms |
| 76L-1). List the diagnosis codes from Q75L. | ______________________________ |
| 76L-2). | ______________________________ |
| 76L-3). | ______________________________ |
| 76L-4). | ______________________________ |
| 76L-5). | ______________________________ |
| 76L-6). | ______________________________ |
| 76L-7). | ______________________________ |
| 76L-8). | ______________________________ |
| 76L-9). | ______________________________ |
| 76L-10). | ______________________________ |
| 77L. Did the shoulder injury occur in the LEFT arm in which a vaccine was injected? | ◯ Yes<br>◯ No<br>◯ Unknown |

| | |
|---|---|
| 77L-1). Please copy and paste the relevant statement from the chart notes on the LEFT shoulder injury. | ______________________ |
| 78L. Did the symptoms of the LEFT shoulder injury begin within the first 7 days following vaccination?<br><br>(Please be sure to use the most complete and reliable information to answer this question when there are discrepancies with more than one source.) | ◯ Yes<br>◯ No<br>◯ No, but increased severity of symptoms<br>◯ Possible<br>◯ Unknown |
| 78L-1). Please copy and paste the relevant statement from the chart notes on the LEFT shoulder injury symptom onset. | ______________________ |
| 79L. Did the symptoms of LEFT shoulder injury persist more than 30 days from the date of vaccination?<br><br>(Please be sure to use the most complete and accurate information to answer this question when there are discrepancies with more than one source.) | ◯ Yes<br>◯ No<br>◯ Possible<br>◯ Unknown |
| 79L-1). Please copy and paste the relevant statement from the chart notes on the LEFT shoulder injury symptom duration. | ______________________ |
| 80L. What was the cause of the LEFT shoulder injury?<br><br>(Please select as many causes as documented.) | ☐ Vaccine<br>☐ Incident (e.g. fall, auto accident)<br>☐ Exercise (e.g. exercise, sports)<br>☐ Daily activity (e.g. overuse, lifting a heavy item, work-related injury, side sleeping)<br>☐ Other medical conditions (e.g. arthritis, chest pain radiating to the shoulder)<br>☐ Unknown (e.g. no explicit cause, insidious/aggravating factors- 'worse with exercise') |
| 80L-1). Please copy and paste the relevant statement from the chart notes on the cause of the LEFT shoulder injury (for all of the above responses). | ______________________ |
| 81L. Does it look like a SIRVA case? | ◯ Yes<br>◯ No<br>◯ Possible<br>◯ Unknown |

#### I. GENERAL COMMENTS

| | |
|---|---|
| 82. Do you have any questions or comments not addressed in the form? Please enter them here. | ______________________ |

### Multimedia Appendix 3:

**Sample extracted temporal expressions:**

- Part of the day: this morning, last night, etc.
    - left shoulder pain after a mechanical ground level fall last evening
- Date: 2/14/17, 10/1, Dec 12, etc.
    - States that her shoulder pain has been an ongoing issue since July 2016
- Weekday: Wednesday, last Monday, yesterday, etc.
    - Was changing his tire on Friday and heard a popping noise in his left shoulder, now has pain with decreased ROM.
- Month: September, etc.
    - left shoulder pain Onset w/ reaching back for seatbelt in Oct
- Season: Spring, Summer, Christmas, etc.
    - I injured my (left) shoulder in a fall last summer (2016)
- Duration: for 3 weeks, x several days, etc.
    - shoulder pn x 2 mo
    - pain, loss of motion left shoulder the past 10 years
    - Who had a fall 2-3 weeks ago - hit the right shoulder against a wall and pain getting worse

Some other temporal data were not extracted because they could not be used to infer the onset timing or duration of the shoulder injury:

- indicates symptom history but without an exact onset date. For example, "patient had been bothered with left shoulder pain"; "shoulder pain (chronic)".
- describes frequency (e.g. "every 4 weeks") or future event (e.g. "next 2 days").

### Multimedia Appendix 4:

**Table S4.1. Types of causes associated with shoulder injuries**

| Order | Type of cause | Description |
|---|---|---|
| 1 | Vaccination | Specific vaccine name or general vaccine terms |
| 2 | Accident | Accidents such as auto accident, fall, hit |
| 3 | Work | Work-related injury |
| 4 | Other medical conditions | Medical conditions that can cause shoulder injury such as arthritis, chest pain radiating to the shoulder |
| 5 | Exercise | Exercise or sports-related injury |
| 6 | Daily activity | Injuries occurred during other daily activities such as lifting groceries, overuse, side sleeping. |
| 7 | Unknown | Insidious or unknown cause |

**Table S4.2. Trigger phrases for identifying causal relationships**

**Trigger phrase before the causal phrase**

*[at above in near] site of*

*[blame* attribute* complain*]*

*[compatible consistent] [for with]*

*[got given getting]*

*[reaction rxn react]*

*[was been]*

*after*

*[associated association] with*

*[because cause due]*

*because of*

*c / w*

*cause of injury*

*caused by*

*complication* of*

*consequence* of*

*doing*

*due to*

*during*

*etiology*

*evidence [for of]*

*except for*

*experienced*

*following*

*from*

*had*

*manifestation of*

*mark* of*

*owing to*

*post*

*receive**

*relate* to*

*result* [from of]*

*s / p*

*[second secondary] to*

*since*

*started with*

*status post*

*status-post*

*was from*

*was given*

[ ] indicates selection of one of the words within the bracket

* indicates capture of all the morphological variants of that term

Example: "Pt presents with a chief complaint of left shoulder pain, no weakness that began 6 months ago after flu shot." In the above example, "after" is the trigger phrase, and "flu shot" is the causal phrase.

**Trigger phrase after the causal phrase**

*causing*

*resulting*

*next [day week]*

*[after later] /word [day week]*

*[day* week*] [after later]*

*was [given administered injected]*

*[reaction rxn react]*

*related*

/word means any word.

Example:

"Received the flu shot on December 13 and the next day she reported she was unable to move her arm."

In the above example, "next day" is the trigger phrase, and "flu shot" is the causal phrase.

**Table S4.3. Terminology for causes of shoulder injury other than vaccination**

**Note:**

Each line represents one entry of the terms

[] indicates selection of one of the words within the bracket

* indicates capture of all the morphological variants of that term

The top-level causes are:

- cause_accident
- cause_work
- cause_other_medical_conditions
- cause_exercise
- cause_daily_activity
- cause_unknown

### cause_accident

abrasion
accident*
accidental fall
airbag
auto accident
bite
blow
car
car accident
CHP
collided
collision
dislocated
dislocation
driving
fall*
FALL:
hit*
incident
landing
landed
mechanical fall
motor vehicle accident
motorcross
motorvehicle accident
motor-vehicle accident

MVA
passenger
punctured
rear ended
rear-end
rearended
rear-ended
separation
slip*
struck
t bone
t boned
t-bone
t-boned
trauma*
traumatic
tripped
twisted
vehicle accident

#### cause_work

job
work
workers compensation

#### cause_other_medical_conditions

abrasion
arithritis
arthritic
arthritis
athritis
back pain
birth
cervical spondylosis
CVA
degenerative
djd
fibromyalgia
mastectomy*
MCA
multifactorial
neck pain
neckpain
neuritis
neuropathy
OA

osteoarthritic
osteoarthritis
polyarthritis
radiating pain
radicular
radiculitis
radiculopathy
referred pain
sciatica
spondyloarthropathy
stroke
surgery
whiplash
whiplashed

#### cause_exercise

aerobics
archer
archery
arena
arrow
athlete
athletics
axel
badminton
ball
baseball
basketball
bat
baton
batter
batting
bench press
biathlon
bicycle
bicycling
bike
biking
billiards
bobsleigh
bocce
boomerang
boules
bow
bowler
bowling

boxer
boxing
bronze medal
bunt
canoe
canoeing
catch
catcher
champion
championship
cleats
club
coach
compete
competing
competition
competitor
crew
cricket
croquet
cross country
curling
cycle
cycling
cyclist
dart
dartboard
deadlifting
decathlon
defense
diamond
discus
dive
diver
diving
dodgeball
doubleheader
dugout
épée
equestrian
fencing
field hockey
fielder
fielding
figure skating
fishing

fitness
football
free throw
frisbee
game*
geocaching
goalie
gold medal
golf
golfer
golfing
got hit
gym
gymnasium
gymnast
gymnastics
hammer throw
hand stand
handball
hang gliding
hardball
helmet
heptathlon
high jump
hitter
hockey
hole-in-one
hoop
horseshoes
huddle
hurdle
ice hockey
ice rink
ice skates
ice skating
infield
infielder
inline skates
inning
jai-alai
javelin
jog
jogger
judo
jump
jump rope

jumper
jumping
karate
kayak
kayaker
kayaking
kickball
kite
kung fu
lacrosse
lawn bowling
league
lift* weight*
long jump
luge
lutz
mallet
martial art*
mitt
offense
ollie
Olympics
orienteering
outfield
outfielder
overhead press
paddle
paddleball
paddling
paintball
parallel bar*
parasailing
parkour
pentathlon
pickleball
ping pong
pitch
pitcher
play*
playground
player
playoffs
pogo stick
pole
pole vault
polo

pool
puck
pull up*
push* up*
quarterback
quiver
race
racer
racewalking
racing
racket
racquetball
rafting
riding
rink
rock climbing
roller-blading
roller blading
rollerblade*
rollerskate*
roller skates
roller skating
roller-skates
roller-skating
row
rower
rowing
rqcquet
rugby
run
runner
running
sailing
scoot*
scuba
scull
sculling
shortstop
shot put
silver medal
skate
skating rink
skeleton
ski
skier
skiing

slalom
sled*
sledder
snorkeling
snowboard
snowboarder
snowboarding
snowshoeing
soccer
softball
somersault
speed skating
sport*
sportsmanship
squash
stadium
strike
sumo wrestling
surf*
surfer
swim*
swimmer
table tennis
taekwondo
taiko
tennis
tetherball
throw
throwing
toboggan
track and field
trampoline
triathlete
triathlon
tricycle
triple jump
triple play
tug of war
ultramarathon
ultramarathoner
umpire
unicycle
unicyclist
vault
vaulter
vaulting

volley
volley ball
volleyball
wakeboarding
water polo
water ski
water skier
water skiing
weightlifter
weightlifting
weight lifting
weight training
weights
wicket
windsurfer
windsurfing
work* out
workout
wrestler
wrestling
yoga

#### cause_daily_activity

carry*
cleaning
grab*
heavy
lift*
massage
overuse*
paint*
pick*
poor posture
reaching
repetitive [injury* injury- injruy injurys injuries injurious injured injure injuring]
repetitive [motion* use* strain* lift* type*]
wake* up

#### cause_unknown

[no none]
[unsure unknown unclear uncertain undertermined unknwon]
[insidious insidiuos incidious]
gradual
[N/A na]

### Multimedia Appendix 5:

**Table S5. Error Analyses on the Validation Dataset**

| |
|---|
| **Error analysis on injury onset** |
| "She has chronic pain--neck, low back, B/L shoulders. She has fibromyalgia, and also fell a few weeks ago which worsened her back pain."<br>NLP incorrectly associated the event ("fall") with the shoulder problem when performing a cross-sentence search. |
| Prior condition reported on Day 0 visit: "My left shoulder pain never went away despite still doing physical therapy and living on NSAIDs. Now it is constant and much worst today."<br>NLP incorrectly captured "today" as the shoulder pain onset date when performing a cross-sentence search. |
| **Error analysis on injury duration** |
| On Day 136, "States in past pain would travel to left shoulder causing numbness to left arm and lasting few days but today denies any numbness."<br>NLP incorrectly identified the injury duration based on a resolved shoulder symptom. |
| **Error analysis on injury cause** |
| "with one day of pain in the left arm and shoulder. denies any injury. Did some lifting yesterday."<br>NLP failed to identify the possible cause (daily activity). |
| "She has been working on the computer a lot. Overhead movement exacerbates the pain...No injury or trauma."<br>NLP failed to identify the possible cause (daily activity). |
| "who complains of left shoulder pain that started 3 weeks ago after vacuuming"<br>NLP failed to identify the possible cause (daily activity). |

"likely subdeltoid bursitis and supraspinatus tendinopathy in the setting of DM likely from acute movement with pain when getting IV placed."

NLP failed to identify the possible cause (accident).

"Patient reports left shoulder pain with movement no trauma. Patient worked for years caring for young children and had to carry and lift them."

NLP identified the cause as 'Unknown', failed to identify the possible cause (daily activity).
